# Comparison of children and young people admitted with SARS-CoV-2 across the UK in the first and second pandemic waves: prospective multicentre observational cohort study

**DOI:** 10.1101/2021.09.14.21263567

**Authors:** Olivia V Swann, Louisa Pollock, Karl A Holden, Alasdair PS Munro, Aisleen Bennett, Thomas C Williams, Lance Turtle, Cameron J Fairfield, Thomas M Drake, Saul N Faust, Ian P Sinha, Damian Roland, Elizabeth Whittaker, Shamez N Ladhani, Jonathan S Nguyen-Van-Tam, Michelle Girvan, Chloe Donohue, Cara Donegan, Rebecca G Spencer, Hayley E Hardwick, Peter JM Openshaw, J Kenneth Baillie, Ewen M Harrison, Annemarie B Docherty, Malcolm G Semple, on behalf of ISARIC4C Investigators

## Abstract

**Background:** Children and young people (CYP) were less affected than adults in the first wave of SARS-CoV-2 in the UK. We test the hypothesis that clinical characteristics of hospitalized CYP with SARS-CoV-2 in the UK second wave would differ from the first due to the combined impact of the alpha variant, school reopening and relaxation of shielding.

**Methods:** Patients <19 years hospitalised in the UK with clinician-reported SARS-CoV-2 were enrolled in a prospective multicentre observational cohort study between 17^th^ January 2020 and 31^st^ January 2021. Minimum follow up time was two weeks. Clinical characteristics were compared between the first (W1) and second wave (W2) of infections.

**Findings:** 2044 CYP aged <19 years were reported from 187 hospitals. 427/2044 (20.6%) had asymptomatic/incidental SARS-CoV-2 infection and were excluded from main analysis. 16.0% (248/1548) of symptomatic CYP were admitted to critical care and 0.8% (12/1504) died. 5.6% (91/1617) of symptomatic CYP had Multisystem Inflammatory Syndrome in Children (MIS-C).

Patients in W2 were significantly older (median age 6.5 years, IQR 0.3-14.9) than W1 (4.0 (0.4-13.6, p 0.015). Fever was more common in W1, otherwise presenting symptoms and comorbidities were similar across waves. After excluding CYP with MIS-C, patients in W2 had lower PEWS at presentation, lower antibiotic use and less respiratory and cardiovascular support compared to W1. There was no change in the proportion of CYP admitted to critical care between W1 and W2.

58.0% (938/1617) of symptomatic CYP had no reported comorbidity. Patients without co-morbidities were younger (42.4%, 398/938, <1 year old), had lower Paediatric Early Warning Scores (PEWS) at presentation, shorter length of hospital stay and received less respiratory support. MIS-C was responsible for a large proportion of critical care admissions, invasive and non-invasive ventilatory support, inotrope and intravenous corticosteroid use in CYP without comorbidities.

**Interpretation:** Severe disease in CYP admitted with symptomatic SARS-CoV-2 in the UK remains rare. One in five CYP in this cohort had asymptomatic/incidental SARS-CoV-2 infection. We found no evidence of increased disease severity in W2 compared with W1.

**Funding:** Short form: National Institute for Health Research, UK Medical Research Council, Wellcome Trust, Department for International Development and the Bill and Melinda Gates Foundation.

Long form: This work is supported by grants from the National Institute for Health Research (award CO-CIN-01) and the Medical Research Council (grant MC_PC_19059) and by the National Institute for Health Research Health Protection Research Unit (NIHR HPRU) in Emerging and Zoonotic Infections at University of Liverpool in partnership with Public Health England (PHE), in collaboration with Liverpool School of Tropical Medicine and the University of Oxford (NIHR award 200907), Wellcome Trust and Department for International Development (215091/Z/18/Z), and the Bill and Melinda Gates Foundation (OPP1209135). Liverpool Experimental Cancer Medicine Centre provided infrastructure support for this research (grant reference: C18616/A25153). JSN-V-T is seconded to the Department of Health and Social Care, England (DHSC). The views expressed are those of the authors and not necessarily those of the DHSC, DID, NIHR, MRC, Wellcome Trust, or PHE.

## Introduction

Children and young people (CYP) were significantly less affected than adults during the first wave of the COVID-19 pandemic, with regards to case numbers, disease severity, hospital admissions and death.^1–4^ The reasons for the predominantly mild disease course in CYP are not yet well defined, although several hypotheses have been proposed, including reduced expression of ACE2 (the binding receptor for SARS-CoV-2) in the lower airways, immunity from prior exposure to seasonal coronaviruses and difference in immune response to acute SARS-CoV-2 infection.^5^ Whilst the clinical profile of CYP with symptomatic SARS-CoV-2 infection (COVID-19) shares similarities with other respiratory viruses^2,3^ (with at-risk cohorts including young infants and those with neurological and cardiac comorbidities^2,4^), the virus also has unusual presentations in the paediatric population. A small proportion of CYP exposed to the virus go on to develop a severe hyperinflammatory syndrome^2^ known as multisystem inflammatory syndrome in children (MIS-C), also known as paediatric inflammatory multisystem syndrome temporally associated with SARS-CoV-2 (PIMS-TS), which shares features of Kawasaki disease and Toxic Shock Syndrome, and often requires management in intensive care.^6,7^

A significant amount of knowledge was gained about the clinical characteristics and outcomes of COVID-19 in CYP during the first wave of the pandemic, however, several external factors changed with the emergence of the second wave. Most UK schools were closed during the first pandemic wave in the spring and summer of 2020, with a few remaining open for children of key workers but were mostly open during the subsequent autumn and winter wave of infection (*Supp Figure A*). This policy reflected the view that the educational, social, health and economic benefits of in-person schooling outweighed the harms associated with school transmission of SARS-CoV-2 at that time. During the first wave, some CYP were identified as extremely clinically vulnerable and advised to shield from all non-essential contact. This advice was removed in autumn 2020. New variants have emerged, including the alpha variant (B.1.1.7) first detected in Southeast England in September 2020, becoming the predominant variant throughout the UK by the end of December.^8,9^ The alpha variant contains mutations that permit some immune escape^10^ in those who had been previously infected, with increased transmissibility^11^, and more severe disease with higher rates of hospitalisation and death in adults.^12^

The emergence of the alpha variant in England also led to concerns of increased transmissibility in CYP as they formed a higher proportion of total cases in England when compared to the first pandemic wave.^13^ This may have been due to the emergence of the variant coinciding with a period when schools were open and subject to increased testing, but the rest of UK society was in “lockdown”.^11^ Whether the alpha variant, dominant in the second wave, causes different symptoms or more severe disease in CYP compared to strains circulating in the first wave has not been analysed in detail.

We test the hypothesis that clinical characteristics of hospitalized children with SARS-CoV-2 in the UK second wave would differ from the first due to the combined impact of the alpha variant, school reopening and relaxation of shielding.

We aimed to characterise and compare the clinical features and outcomes of CYP aged <19 years who were hospitalised with SARS-CoV-2 infection during the first and second waves across England, Scotland, and Wales from 17^th^ January 2020 to 31^st^ January 2021, as part of the International Severe Acute Respiratory and Emerging Infection Consortium -World Health Organisation, Clinical Characterisation Protocol in the United Kingdom (ISARIC WHO CCP-UK).

## Methods

### Study design, setting and participants

The protocol, associated documents, and details of the Independent Data and Material Access Committee (IDAMAC) are available at https://isaric4c.net.^14^ We included all patients aged <19 years with clinician-reported SARS-CoV-2 infection who were enrolled into the ongoing, prospective ISARIC WHO CCP-UK cohort study involving National Health Service (NHS) hospitals in England, Wales, and Scotland between 17^th^ January 2020 to 31^st^ January 2021 who had at least two weeks of outcome data available (Wave 1 - 17^th^ January 2020 – 31^st^ July 2020, Wave 2 - 1^st^ August 2020 – 31^st^ January 2021).^15^ Patients were managed by their local clinicians and participation in this study had no influence on management. We used the Strengthening the Reporting of Observational Studies in Epidemiology (STROBE) guidelines for reporting this observational study.

### Data collection

Data were collected from healthcare records onto case report forms through a secure online database, REDCap (Research Electronic Data Capture, Vanderbilt University, hosted by the University of Oxford, UK). Demographic (including age, sex, self-reported ethnicity, postal code) and baseline data (including comorbidities and regular medications taken) alongside data on symptoms, clinical signs, laboratory and pathology investigations, and treatments received while admitted were collected. Centres also recorded whether their team had treated patients as having MIS-C.

### Clinician-reported SARS-CoV-2

Patients were included in this report if the study team had reported them as “laboratory-proven” SARS-CoV-2. Where patients were reported as “suspected” SARS-CoV-2, the patients’ virological data were reviewed, and patients were included if there was documented evidence of a positive PCR, serology or lateral-flow testing for SARS-CoV-2.

### “Incidental SARS-CoV-2” and “other reason for admission” variables

We reviewed all available free text for evidence of incidental SARS-CoV-2 infection (e.g., hospitalisation for elective surgery, road traffic accidents, or drug overdoses, see *Supplementary Methods*). Patients in whom SARS-CoV-2 was judged to be incidental or who were asymptomatic at the time of assessment for SARS-CoV-2 were censored from the main analysis.

### Symptomatic patients with SARS-CoV-2

Patients who had reported symptoms together with those missing symptom data are referred to as “symptomatic CYP,” however, it is possible that not all symptoms were due to SARS-CoV-2.

### Outcomes

The primary outcomes of this study were admission to critical care (high dependency units (HDUs) or paediatric intensive care units (PICUs)), development of MIS-C, and in-hospital mortality for CYP and young people with symptomatic SARS-CoV-2 infection (COVID-19). We also examined the need for any respiratory and cardiovascular (inotropic) support.

### Bias

As specialist children’s hospitals (tertiary centres) are more likely to have both paediatric critical care facilities and paediatric research teams with capacity to support participating in the study, it is possible that CYP admitted to these centres are over-represented. We compared the proportion of CYP who were reported from hospitals with onsite access to a PICU to ascertain whether this differed between the waves, potentially influencing the severity of patients reported (see *Supplementary Results*).

To explore how well the ISARIC data reflected regional variations in SARS-CoV-2 prevalence, we also compared the number of CYP reported to ISARIC against the numbers of local SARS-CoV-2 cases identified by pillar 1 (hospital) and pillar 2 (community) testing across NHS regions (see *Supplementary Results*).

Second wave data may have a lower proportion of patients with MIS-C because the condition typically develops 2 to 4 weeks after the initial infection. As data collection for this analysis ended just before the end of the second wave this could bias towards fewer severely ill CYP in the second wave. To reduce this bias, this patient group was censored when comparing clinical characteristics between waves.

### Statistical analysis

Continuous variables are displayed as means (standard deviations) or if non-normally distributed as medians (interquartile ranges). Categorical variables are presented as frequencies (percentages) unless otherwise stated. For univariable comparisons, we used Welch’s *t*, analysis of variance, Mann-Whitney U, or Kruskal-Wallis tests, according to data distribution. We compared categorical data by using χ^2^ tests and considered a p value below 0.05 to be statistically significant. All tests were two sided and we made no adjustment for multiple comparisons. A directed acyclic graph was constructed prior to undertaking a mixed effect multivariable analysis (*Supp Figure B*). Hospital was included as a random effect in the multivariable analysis. Parsimonious criterion-based model building used the following principles: relevant explanatory variables were identified from previous studies; interactions were checked at first order level; final model selection was informed by the Akaike information criterion and C statistic, with appropriate assumptions checked including the distribution of residuals. We used R (R Core Team version 3.6.3, Vienna, Austria) for statistical analyses, with packages including tidyverse, finalfit lubridate, UpSetR and ggplot2.

### Patient and public involvement

Patients and the public were not involved in the design, conduct, or reporting of this rapid response research which is part of an ongoing urgent public health research study, however their involvement is in now progress.

### Legal basis for data collection and ethics approval

In England and Wales routine anonymised data from medical records was collected without the need for consent under regulation 3 (4) of the Health Service (Control of Patient Information) Regulations 2002. In Scotland, a waiver of need for consent was obtained from the Public Benefit and Privacy Panel. Ethical approval was given by the South Central–Oxford C Research Ethics Committee in England (reference 13/SC/0149) and the Scotland A Research Ethics Committee (reference 20/SS/0028).

## Results

Between 17^th^ January 2020 and 31^st^ January 2021, 187,267 admissions of all ages were enrolled. There were 2044 (1.1%) CYP aged <19 years of age with clinician-reported SARS-CoV-2 infection reported from 187 hospitals across England, Scotland, and Wales. Of these, 1,540 (75.3%) had symptoms at presentation, 427 (20.6%) had asymptomatic or incidental SARS-CoV-2 infection and 77 (3.8%) were missing data on symptoms (*Figure 1*). Of the symptomatic CYP, 91 were identified as having MIS-C.

**Figure 1.**
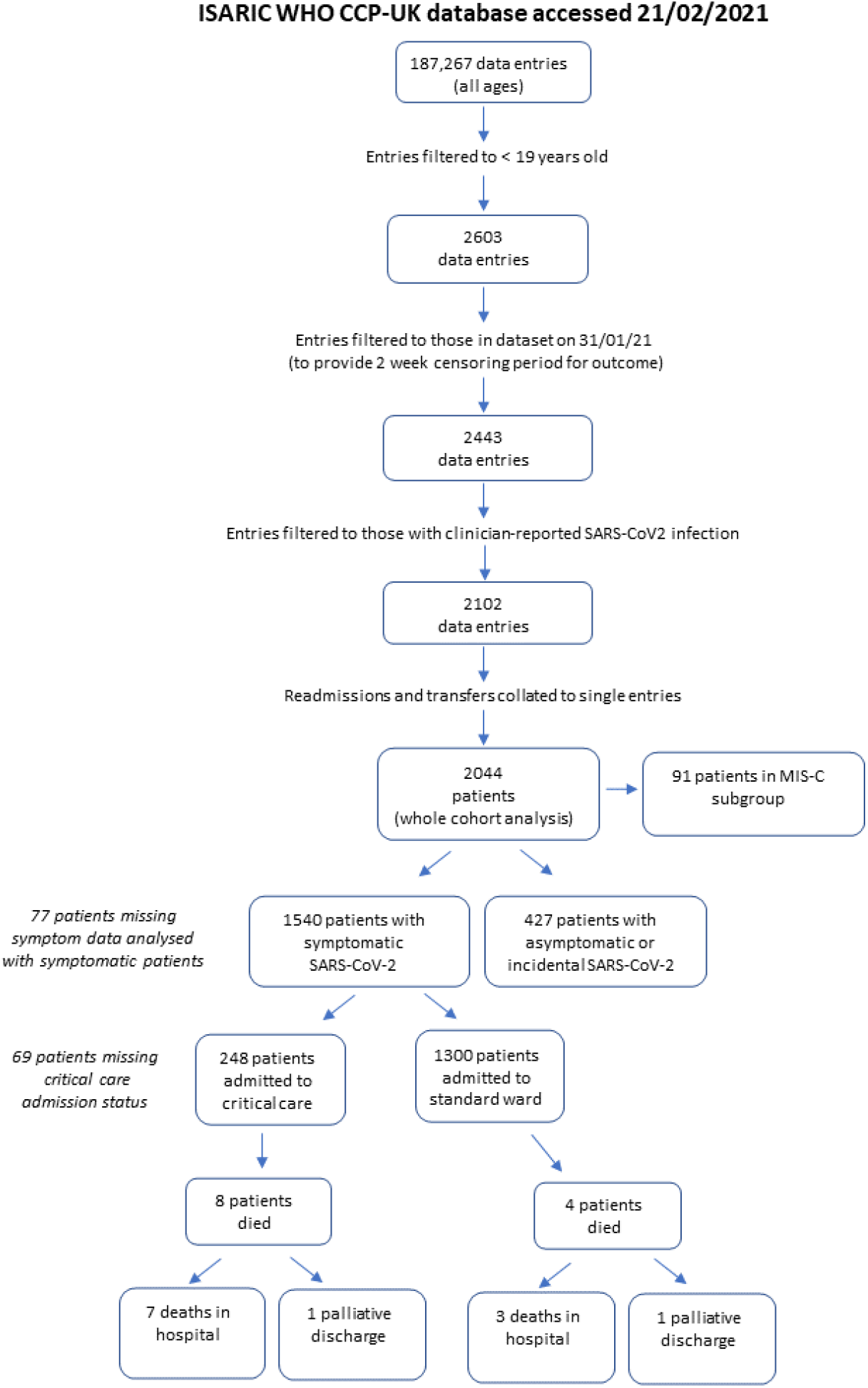
Flowchart of patient inclusion and outcomes

### Demographics of symptomatic CYP admitted in the first wave vs second wave

In total, 764 CYP were admitted during wave one (W1, 17^th^ January to 31st July 2020) and 1,280 during wave two (W2, 1^st^ August 2020 to 31st January 2021). CYP in W2 were significantly older (median age 6.5 years, IQR 0.3 - 14.9) than W1 (4.0 (0.4 - 13.6), p = 0.015) *(Table 1)*. CYP of South Asian ethnicity were over-represented in W2 (19.1%, 155/810) compared to W1 (13.6%, 78/575, p = 0.008). W2 saw a lower proportion of likely hospital-acquired SARS-CoV-2 (2.2% (21/952) vs W1, 6.9% (46/665), p <0.001). Fever was more common in W1 (76.8% (491/639) vs 63.6% (544/855), p <0.001, *Supp Table B*), otherwise presenting symptoms were very similar (*Supp Figure E*). Comorbidities were similar in W2 and W1 for symptomatic CYP (*Supp Table C*) and the whole cohort (*Supp Table D*).

**Table 1.** Demographics of patients <19 years by wave of SARS-CoV-2 pandemic (excluding patients with asymptomatic and incidental SARS-CoV-2). First wave ending 31^st^ July 2020. Values are numbers (percentages) unless stated otherwise. IQR = interquartile range. IMD = Indices of multiple deprivation.

|  | Total N |  | First | Second | p |
| --- | --- | --- | --- | --- | --- |
| Total N (%) |  |  | 665 (41.1) | 952 (58.9) |  |
| Age at assessment (Years) | 1617 (100.0) | Median (IQR) | 4.0 (0.4 to 13.6) | 6.5 (0.3 to 14.9) | 0.015 |
| Age | 1617 (100.0) | <1 mth | 48 (7.2) | 74 (7.8) | 0.011 |
|  |  | >1mth <1 y | 181 (27.2) | 226 (23.7) |  |
|  |  | 1-4 y | 117 (17.6) | 139 (14.6) |  |
|  |  | 5-9 y | 90 (13.5) | 101 (10.6) |  |
|  |  | 10-14 y | 102 (15.3) | 182 (19.1) |  |
|  |  | 15-19 y | 127 (19.1) | 230 (24.2) |  |
| Sex at Birth | 1613 (99.8) | Male | 363 (54.6) | 500 (52.5) | 0.463 |
|  |  | Female | 301 (45.3) | 449 (47.2) |  |
|  |  | (Missing) | 1 (0.2) | 3 (0.3) |  |
| Ethnicity | 1385 (85.7) | White | 330 (49.6) | 472 (49.6) | 0.008 |
|  |  | Black | 49 (7.4) | 56 (5.9) |  |
|  |  | South Asian | 78 (11.7) | 155 (16.3) |  |
|  |  | Other ethnic minority | 118 (17.7) | 127 (13.3) |  |
|  |  | (Missing) | 90 (13.5) | 142 (14.9) |  |
| IMD quintile | 1491 (92.2) | 1 (most deprived) | 212 (31.9) | 330 (34.7) | 0.224 |
|  |  | 2 | 130 (19.5) | 180 (18.9) |  |
|  |  | 3 | 87 (13.1) | 144 (15.1) |  |
|  |  | 4 | 82 (12.3) | 113 (11.9) |  |
|  |  | 5 (least deprived) | 101 (15.2) | 112 (11.8) |  |
|  |  | (Missing) | 53 (8.0) | 73 (7.7) |  |
| Potential hospital acquired SARS-CoV-2 | 1617 (100.0) | No | 619 (93.1) | 931 (97.8) | <0.001 |
|  |  | Yes | 46 (6.9) | 21 (2.2) |  |
| Any comorbidity | 1617 (100.0) | No/Unknown | 367 (55.2) | 571 (60.0) | 0.062 |
|  |  | Yes | 298 (44.8) | 381 (40.0) |  |

### Severity at presentation, treatments received and outcomes in symptomatic CYP (excluding MIS-C) examined by wave

Paediatric Early Warning Score (PEWS) at presentation was lower in W2 than W1, with 41.2% (343/832) of CYP in W2 having a PEWS >2 at presentation vs 48.9% in W1 (291/595, p = 0.005, *Table 2* and *Supp Figure F*). Median length of stay was very short at 2 days (IQR 1-4) for both waves (*Supp Figure G*). We found no difference in the proportion of symptomatic CYP admitted to critical care in W1 vs W2 (12.9% (78/604) vs (12.7% (109/855, p = 0.989, *Table 2*). CYP in W2 had lower antibiotic use than W1 (58.0% (467/806) vs 70.6% (415/588, p <0.001)), were less likely to receive high flow oxygen, non-invasive or invasive respiratory support as well as fewer patients in W2 requiring inotropic support (1.0% (8/780) vs 3.6% (21/580, p = 0.002)). CYP in W2 were more likely to receive oral steroids. These associations persisted in a sensitivity analysis of the whole cohort (*Supp Table F*).

**Table 2.**
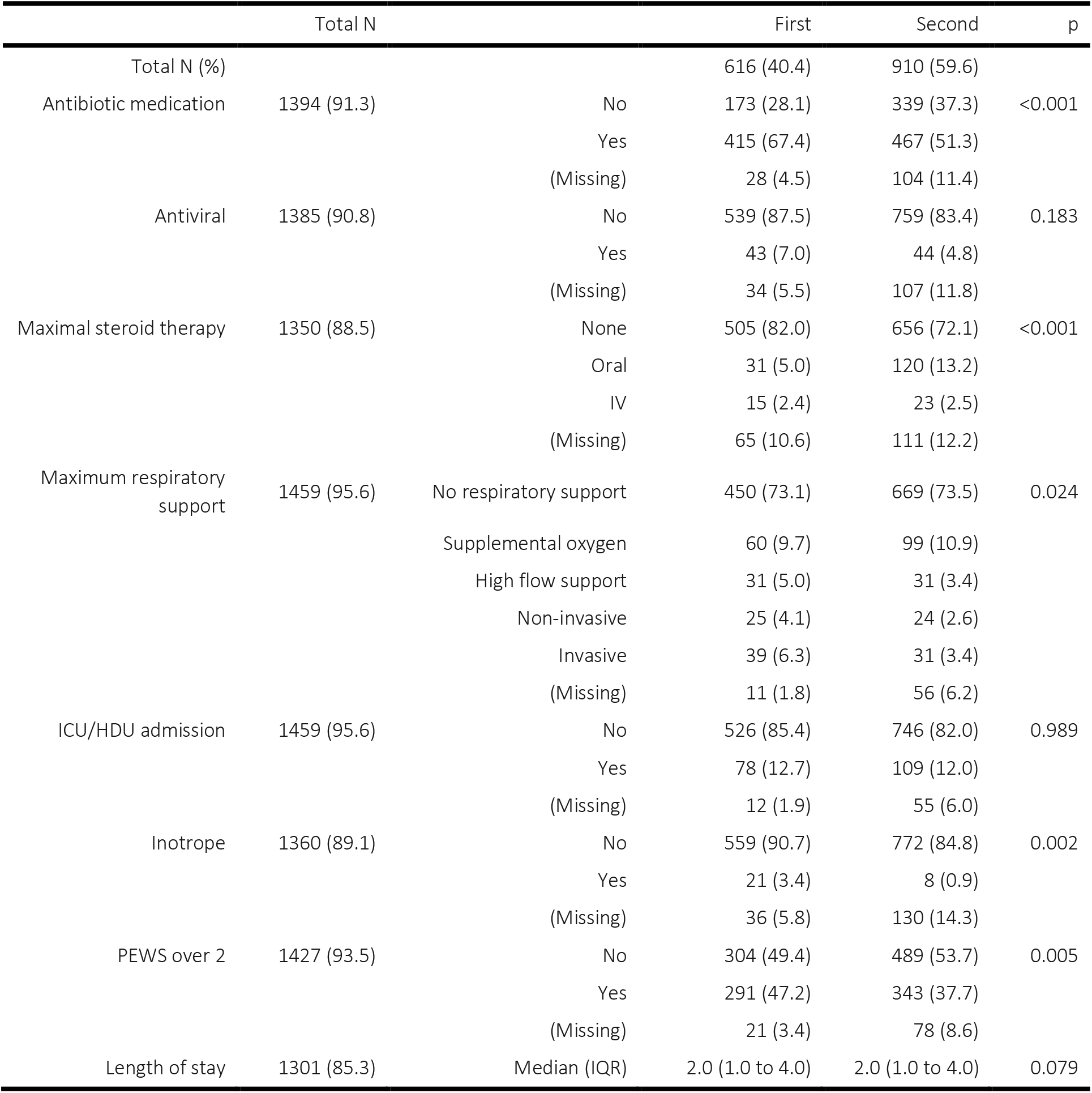
Comparison of treatments received and outcomes by wave (excluding asymptomatic and incidental SARS-CoV-2 infections and patients with Multisystem Inflammatory Syndrome in Children (MIS-C)). IQR = interquartile range. ICU = intensive care unit, HDU = high dependency unit. PEWS = Paediatric Early Warning Score.

### CYP with MIS-C

There were 163 potential MIS-C patients, of whom 91 were confirmed by sites, 46 had other diagnoses and there was no response for 26 patients (*Supp Figure H*). There was no significant difference in PEWS at presentation for patients with MIS-C between W1 and W2, but length of stay was shorter in W2 compared to W1 (median 6.0 days (4.0 to 10.0) vs 8.5 (5.8 to 12.0, p = 0.031). CYP with MIS-C in W2 were less likely to receive IVIg than in W1 (59.5% (25/42) vs 83.7% (41/49), p = 0.018, *Supp Table G*), but there was no difference in use of antibiotics, steroids (oral or IV), immunomodulators, respiratory or cardiac support or critical care admission.

### Factors associated with critical care admission

We reviewed the demographics and key clinical characteristics of CYP admitted to critical care, excluding those with asymptomatic or incidental SARS-CoV-2 infection (but including those with MIS-C). On univariable analysis, age groups 5-9 and 10-14 years were associated with critical care admission, as was non-white ethnicity, hospital-acquired SARS-CoV-2 infection, PEWS >2 at admission and presence of an underlying comorbidity (*Supp Table H*). Of the 248 children admitted to critical care, 58.9% (146/248) were aged ≤ 11 years, i.e., in an age group with no current licenced vaccine available. On detailed review, comorbidities associated with critical care admission included prematurity, neurological comorbidity, neurodisability, respiratory comorbidity (excluding asthma) and cardiac comorbidities (*Supp Table I*). Neurological comorbidity was often present in addition to one or more additional comorbidities (*Supp Figure I*). Whilst the majority of CYP admitted to critical care with symptomatic SARS-CoV-2 had comorbidities, 45.2% (112/248) had no reported comorbidity.

### CYP in W2 were no more likely to be admitted to critical care than W1 after excluding MIS-C

As our analysis period likely underestimates the proportion of CYP with MIS-C in W2 (see *Methods*), these patients were excluded from the multivariable analysis to reduce bias when comparing severity between the waves. Neonates and CYP aged 10-14 years and 15-19 years were more likely to be admitted to critical care (*Figure 2* and *Supp Table J*). Other ethnic minorities (i.e., not white, black or South-Asian) were significantly associated with critical care admission as was the presence of one or more comorbidities, and a PEWS of ≥2 at presentation. No association was seen between indices of multiple deprivation (IMD) or sex and admission to critical care. After taking patient demographics, comorbidity count, and PEWS score at presentation into account, we found that CYP were no more likely to be admitted to critical care in W2 when compared to W1.

**Figure 2.**
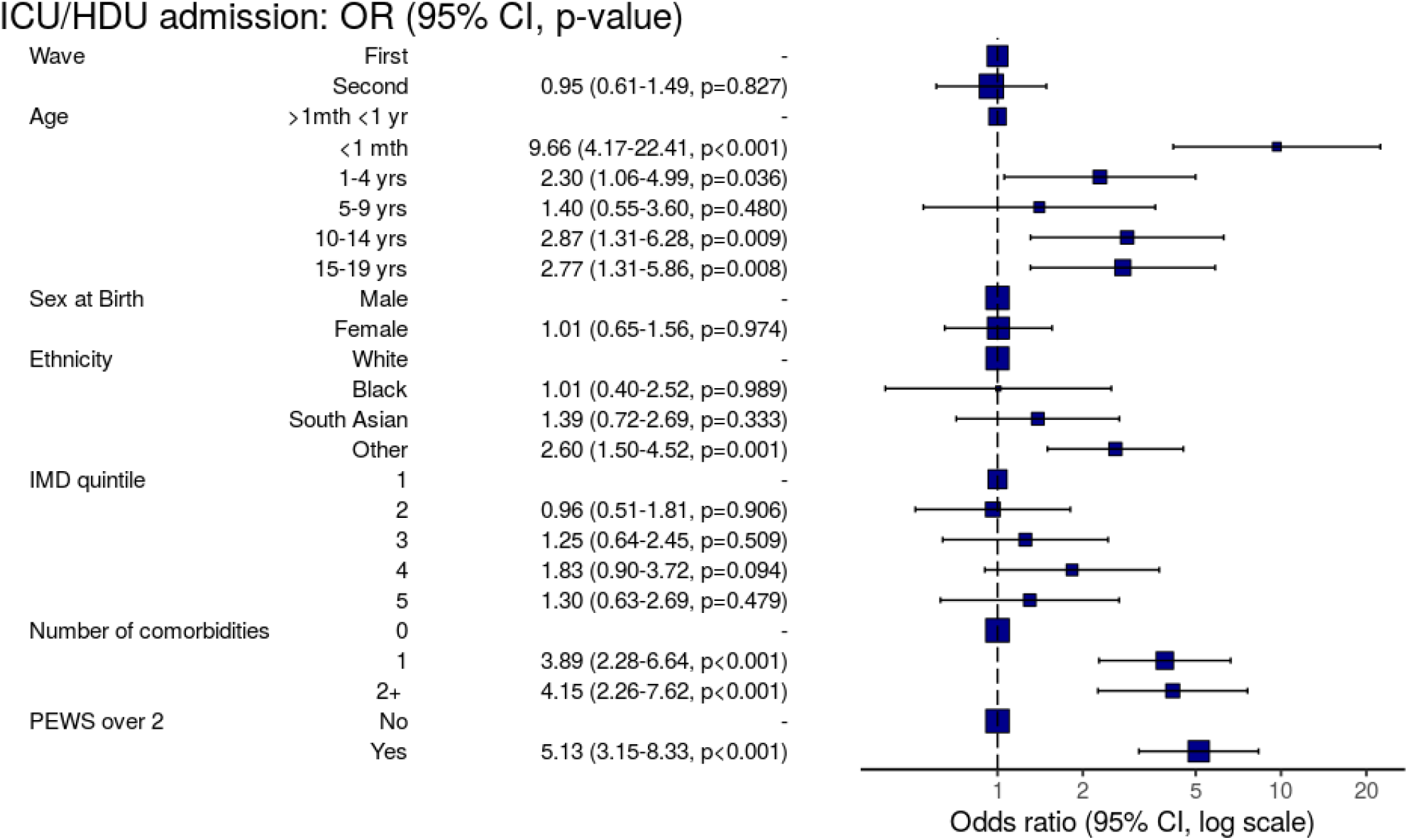
Forest plot of factors associated with admission to critical care unit (excluding asymptomatic and incidental SARS-CoV-2 infections and patients with Multisystem Inflammatory Syndrome in Children (MIS-C)). Other = Other ethnic minorities. IMD = Indices of multiple deprivation (1 = most deprived, 5 = least deprived). PEWS = Paediatric early warning score at presentation. CI = 95% confidence interval.

### Factors associated with death

Outcome data were available for 1504/1617 symptomatic CYP where we identified 12 deaths (10 in hospital and 2 palliative discharges) and an overall mortality rate of 0.8% (12/1504). Information was available for 11 of these deaths. All 11 had significant comorbidities (severe neurodisability, malignancy, very premature, complex congenital heart disease, bacterial sepsis and complex life-limiting comorbidities). Five were < 5 years old and six were > 15 years old.

### Symptomatic patients with and without reported comorbidities (including those with MIS-C)

Patients without a reported comorbidity made up 58.0% (938/1617) of the symptomatic cohort (*Supp Table K*). Of CYP without a reported comorbidity, 70.4% (660/938) were ≤ 11 years (i.e. in an age group with no current licenced vaccine available) and 42.4% (398/938) were aged under 1 year. Among CYP ≥ 12 years, 47.5% (278/585) had no comorbidity recorded on admission. Patients without reported comorbidities had a lower PEWS score at presentation than those with comorbidities (2.0 (1.0-4.0) vs 2.0 (1.0 – 5.0), p = 0.014), and a shorter length of stay (2.0 (1.0-3.0) vs 3.0 (1.0-7.0), p<0.001). CYP with no reported comorbidities were less likely to receive antiviral therapy, steroids, and all forms of respiratory support than in CYP with comorbidities (*Table 3*). Whilst the majority of CYP with no reported comorbidities received ward-level care, 12.9% (112/876) were admitted to critical care, however only 6.6% (58/876) required invasive or non-invasive ventilation. A sensitivity analysis showed that the rates of critical care admission, invasive and non-invasive ventilation, IV steroids and inotropes in CYP without reported comorbidities were driven by the subgroup with MIS-C (*Supp Table L*).

**Table 3.** Treatments received stratified by comorbidity (patients with asymptomatic or incidental SARS-CoV-2 infections excluded). ICU = intensive care unit. HDU = high dependency unit. PEWS = Paediatric Early Warning Score at presentation.

|  | Total N |  | No/Unknown Comorbidity | Comorbidity present | p |
| --- | --- | --- | --- | --- | --- |
| Total N (%) |  |  | 938 (58.0) | 679 (42.0) |  |
| Antibiotic medication | 1477 (91.3) | No | 301 (32.1) | 216 (31.8) | 0.273 |
|  |  | Yes | 529 (56.4) | 431 (63.5) |  |
|  |  | (Missing) | 108 (11.5) | 32 (4.7) |  |
| Antiviral | 1476 (91.3) | No | 787 (83.9) | 595 (87.6) | 0.027 |
|  |  | Yes | 42 (4.5) | 52 (7.7) |  |
|  |  | (Missing) | 109 (11.6) | 32 (4.7) |  |
| Maximal steroid therapy | 1422 (87.9) | None | 713 (76.0) | 475 (70.0) | <0.001 |
|  |  | Oral | 53 (5.7) | 103 (15.2) |  |
|  |  | IV | 42 (4.5) | 36 (5.3) |  |
|  |  | (Missing) | 130 (13.9) | 65 (9.6) |  |
| Maximum respiratory support | 1550 (95.9) | No respiratory support | 738 (78.7) | 427 (62.9) | <0.001 |
|  |  | Supplemental oxygen | 59 (6.3) | 108 (15.9) |  |
|  |  | High flow support | 21 (2.2) | 47 (6.9) |  |
|  |  | Non-invasive | 25 (2.7) | 35 (5.2) |  |
|  |  | Invasive | 33 (3.5) | 57 (8.4) |  |
|  |  | (Missing) | 62 (6.6) | 5 (0.7) |  |
| ICU/HDU admission | 1548 (95.7) | No | 764 (81.4) | 536 (78.9) | <0.001 |
|  |  | Yes | 112 (11.9) | 136 (20.0) |  |
|  |  | (Missing) | 62 (6.6) | 7 (1.0) |  |
| Inotrope | 1451 (89.7) | No | 764 (81.4) | 612 (90.1) | 0.274 |
|  |  | Yes | 47 (5.0) | 28 (4.1) |  |
|  |  | (Missing) | 127 (13.5) | 39 (5.7) |  |
| Total PEWS | 1518 (93.9) | Median (IQR) | 2.0 (1.0 to 4.0) | 2.0 (1.0 to 5.0) | 0.014 |
| PEWS over 2 | 1518 (93.9) | No | 471 (50.2) | 351 (51.7) | 0.315 |
|  |  | Yes | 380 (40.5) | 316 (46.5) |  |
|  |  | (Missing) | 87 (9.3) | 12 (1.8) |  |
| Length of stay | 1368 (84.6) | Median (IQR) | 2.0 (1.0 to 3.0) | 3.0 (1.0 to 7.0) | <0.001 |

### Asymptomatic and incidental SARS-CoV-2

We observed a significant increase in the proportion of patients who were asymptomatic at the time of SARS-CoV-2 detection from 10.4% (78/751) in the first wave to 24.7% (300/1214, p <0.001) in the second wave (*Supp Figure E*). CYP with asymptomatic or incidental SARS-CoV-2 were older (median age 11.2 years (IQR 1.5 - 15.9) vs 5.3 years (IQR 0.4 - 14.2, p <0.001, *Supp Table M*), more likely to have hospital-acquired SARS-CoV-2 infection (12.4% (53/427) vs 4.2% (65/1540), p <0.001), have a reported comorbidity (51.3% (219/427) vs 43.7% (673/1540), p = 0.006) and evidence of an alternative reason for admission (see *Supp Methods*, 30.9% (132/427) vs 5.3% (82/1540, p <0.001). They also had a lower median PEWS on presentation (1.0 (IQR 0.0 - 2.0) vs 2.0 (IQR 1.0 to 4.0), p <0.001). No differences were seen in sex or ethnicity or IMD (*Supp Table N*).

## Discussion

The ISARIC WHO CCP-UK study recruited 2044 CYP with SARS-CoV-2 between 17^th^ January 2020 and 31^st^ January 2021 of whom 20.6 % (427/2044) had asymptomatic or incidental SARS-CoV-2 infections. Of the symptomatic CYP, 5.6% (91/1617) had MIS-C and 16.0% (248/1548) were admitted to critical care. Within the symptomatic group, 0.8% (12/1504) died. There were 665 symptomatic CYP in W1 and 952 in W2, with those in W2 being older, more likely to be of South Asian ethnicity and with a lower proportion of hospital acquired SARS-CoV-2. Reassuringly, there was no evidence of a negative impact of relaxation in guidance to “shield” vulnerable CYP in the latter part of 2020,^16^ with similar prevalence of comorbidities across the two waves.

Despite concerns about more severe disease and fatalities associated with the alpha variant in adults, relaxation of shielding advice and increases in face-to-face schooling, no difference was found in the proportion of symptomatic CYP admitted to critical care between the two waves, instead pointing to less severe disease in W2. After excluding patients with MIS-C, CYP admitted during W2 had a lower PEWS at presentation, lower antibiotic use and less respiratory and cardiovascular support compared to W1. Oral steroid use was higher in W2, likely because of changes in national guidance adopting the results of the RECOVERY trial.^17^ Whilst there was no difference in PEWS at presentation, respiratory or cardiovascular support in CYP with MIS-C between the waves, those in W2 were less likely to receive IVIg. This may reflect increases in clinician confidence, local guidance, or decreased availability of the therapy.

After exclusion of CYP with MIS-C, factors associated with admission to critical care remained very similar to our first report,^2^ with neonates and ages 10-14 and 15-19 years associated with admission in addition to ethnicity, PEWS at presentation and number of comorbidities.

Of particular interest were CYP without comorbidities. Those under 1 year comprised 42.4% of CYP admitted without comorbidities - an age group commonly admitted for brief periods of observation for viral illnesses. CYP with no reported comorbidities had a lower PEWS at presentation, shorter length of stay and received less respiratory support. However, 12.9% (112/876) of all hospitalised CYP without comorbidities were admitted to critical care, with 53 of these having MIS-C. CYP with MIS-C were also responsible for much of the invasive and non-invasive ventilation, inotrope use and IV steroids in the CYP group without comorbidities. Overall, the majority of children in hospital with symptomatic infection had no reported comorbidities (58.0% (938/1617). Of these CYP without reported comorbidities, 70.4% (660/938) were ≤ 11 years, representing a significant group in whom there is no current licenced vaccine available, while 47.5% (278/585) of CYP of vaccine-licensed age had no reported comorbidities. This suggests that targeting prevention strategies, such as vaccines, on the basis of comorbid status or risk groups will have somewhat limited impact on hospitalisation in CYP of vaccine licensed age and overall.

The ISARIC WHO CCP-UK study provides an extensive, detailed and prospective dataset, continuously collected since the start of the pandemic and is uniquely placed to monitor changes in the characteristics of hospitalised CYP with SARS-CoV-2 in the UK from emergence to evolution of the pandemic. Unlike routine and population-based studies, data collection for ISARIC is focused on characterisation of novel emerging disease. Due to the need to retain a “lite” data collection process, some data terms are not relevant to CYP. Reporting to ISARIC is voluntary and is likely to lead an underestimate in the number of asymptomatic or incidental cases reported, as some sites may have chosen only to report “true” COVID-19 cases. Genotype data was not available.

It is important to recall that the study definition of critical care is not just intensive care but includes high dependency units in secondary care centres. In paediatrics, patients are often admitted to these wards for close observation, without intensive therapy. This is borne out by our study finding much lower rates of high-level respiratory or cardiovascular support than rates of critical care admission.

Curtailment of analysis on 31^st^ January 2021, after the peak of the second wave and with community cases falling, was planned due to the urgent need to provide paediatric data on the alpha variant to inform public health policy at that time. However, MIS-C typically presents 2-4 weeks after infection, and therefore cases of MIS-C due to infection acquired in the second wave will be underestimated.

A major strength of our study is that it highlights the importance of differentiating whether CYP are hospitalised because of COVID-19 i.e., disease caused by SARS-CoV-2 infection or with SARS-CoV-2 infection that was incidental. Analyses performed without excluding CYP where SARS-CoV-2 is incidental will produce misleading results. Describing and identifying factors predicting severity of COVID-19 could be biased in either direction by inclusion of both a high proportion of asymptomatic, or incidental infections, and inclusion of CYP admitted to critical care for unrelated reasons such as trauma.

Data collection before and after the emergence of the alpha variant provides reassuring evidence that clinical characteristics in CYP did not change over time coincident with the rise to dominance of this strain. There is evidence in adults that the alpha variant is not only associated with higher transmissibility, but also higher risk of hospital admission^18^ and death,^12,19,20^ although estimates of case-fatality rates may be limited by confounding factors.^19^ However, most studies did not include CYP, and reported risks appear to be age dependent. In a large community-based UK retrospective cohort, Nyberg et al. found an increased risk of hospital admission and mortality in adults older than 30 years testing positive for the alpha variant, but found no difference for young people under 20 years.^18^ In a brief report, Brookman et al. compared characteristics of 20 CYP admitted to a single London hospital in W1 to 60 CYP in W2 with no difference in demographics or increase in severity of disease.^21^ Our larger and more detailed study supports these findings.

Much of the focus of recent reports in SARS-CoV-2 in CYP aims to identify comorbidities associated with critical care admission,^22,23^ particularly in the discussion about vaccination of CYP. The granularity of our study allowed us to also examine CYP admitted without comorbidities in detail. While this group appears to be driven by infants with short hospital stays for brief observation, 12.9% of CYP admitted without comorbidities required critical care admission. CYP with MIS-C made up half of this group, but this also suggests that there may still be a group of previously well CYP with as yet unclear risk factors for critical care.

Understanding of asymptomatic SARS-CoV-2 infection has increased throughout the pandemic, particularly in CYP and it is clear that to be clinically useful, studies must identify CYP who have asymptomatic or incidental infection among those who are hospitalised. Two single-centre studies from the USA have reported that between 40-45% of their paediatric admissions with SARS-CoV-2 were either incidental or unlikely to be due to the virus itself.^24,25^ Brookman et al. reported a prevalence of asymptomatic/incidental infection in their cohort of 33%.^21^ Our finding that at least 21% of reported cases in our cohort were asymptomatic/incidental may be an underestimate given that reporting was voluntary and the variable use of free text to record these details.

Differences in demographics and symptomatology between W1 and W2 may reflect changes in testing patterns, infection control and surveillance practices over the course of the pandemic.^26^ UK hospitals gradually moved from testing based on case definition (with key symptoms of fever, cough, respiratory distress, or loss of sense of taste/smell) to universal testing of all admissions. Reduction in hospital-acquired infection may be due to improved infection control procedures, earlier detection of community acquired infection, or both.

Accepting the limitations above, we provide evidence suggesting the emergence of the alpha variant did not lead to more severe disease in CYP in the UK. With the Delta variant now dominating in the UK, our study serves an exemplar of both the strengths and limitations of large hospital-based studies in informing immediate public health approaches to emerging new variants. The key strength of our study is in providing a granularity of individual patient data which allows us to look in detail at clinical presentations and outcomes, identify important sources of bias, and provide comprehensive data over time. The key limitation is that this nuanced approach takes time to perform and is outpaced by the rapid evolution of this pandemic. As a result, initial incomplete data is by necessity used to inform policy, while more accurate information may only be gained in retrospect.

We urge other paediatric cohort studies to develop processes to define and record asymptomatic and incidental SARS-CoV-2 infection and differentiate this in analyses from COVID-19 disease. In addition, this study raises the possibility of as yet unidentified risk factors for critical care in CYP without comorbidities. As new variants of SARS-CoV-2 emerge, there is no guarantee that the generally mild disease observed in CYP to date will continue to predominate. Paediatricians and epidemiologists must remain vigilant in monitoring patterns of SARS-Cov-2 infection in CYP and develop more efficient systems to inform policy and clinical practice with speed and accuracy.

## Supporting information

Strengthening the Reporting of Observational Studies in Epidemiology

## Data Availability

Data are available for reuse through a secure data sharing platform. Access is welcome through the ISARIC4C Independent Data and Material Access Committee (https://isaric4c.net).

https://isaric4c.net/sample_access/

## Acknowledgements

This work uses data provided by patients and collected by the NHS as part of their care and support #DataSavesLives. We are extremely grateful to the 2,648 frontline NHS clinical and research staff and volunteer medical students, who collected this data in challenging circumstances; and the generosity of the participants and their families for their individual contributions in these difficult times.

## Transparency

The senior author (the manuscript’s guarantor) affirms that the manuscript is an honest, accurate, and transparent account of the study being reported; that no important aspects of the study have been omitted; and that any discrepancies from the study as planned (and, if relevant, registered) have been explained.

## Dissemination plans

Dissemination to participants and related patient and public communities: ISARIC4C has a public facing website [ISARIC4C.net] and twitter account (@CCPUKstudy). We are engaging with print and internet press, television, radio, news, and documentary programme makers.

## Structured author contributor role taxonomy (CRediT)

Concept: OVS, KAH, JSN-V-T, PJMO, JKB, MGS. Methodology: OVS, LP, JKB, EMH, ABD, MGS. Software: OVS, CJF, TMD. Validation: LP, KAH, MGS. Analysis: OVS, LP, AB, CJF, TMD, DR, EMH, ABD, MGS. Investigation: OVS, KAH, JKB, EMH, ABD, MGS. Resources: PJMO, JKB, MGS. Data curation: OVS, LP, KAH, AB, TCW, LT, EMH. Writing - original draft: OVS, LP, KAH, APSM, ABD, MGS. Writing - Review and Editing: OVS, LP, KAH, APSM, AB, TCW, LT, CJF, TMD, SNF, IPS, DR, EW, SNL, JSN-V-T, PJMO, JKB, EMH, ABD, MGS. Visualisation: OVS, CJF, TMD. Supervision: MGS. Administration: KAH, MG, ChD, CaD, RGS, HEH. Funding: PJMO, JKB, ABD, MGS.

## Supplementary Information

### Supplementary Methods

Data on clinical progress were collected on days 1, 3, 6 and 9 of admission and, if applicable, on the day of admission to critical care, as were data on interim outcome status at day 28 and final outcome (discharged alive/palliative discharge/in-hospital death) when that occurred.

#### Variables

Baseline vital signs were used to calculate a Paediatric Early Warning Score (PEWS) which provided a measure of severity of illness at presentation.^1^ In addition to the variables used for comorbidities in our original report, we also included a neurodevelopmental comorbidity category which included children and young people (CYP) with learning disability, autism spectrum disorders and attention deficit hyperactivity disorder.^2^ Oxygen delivered by high-flow nasal cannulae was assigned as “high flow support” which is available in critical care environments and on some wards. CYP admitted for >5 days before testing for SARS-CoV-2 infection were categorised as potential hospital acquired infection.

#### Length of stay

Length of stay was calculated from date of assessment in hospital for SARS-CoV-2 infection and date of discharge (where this was recorded) and was available for patients who had a recorded discharge by 28 days.

#### Age

Age was calculated based on date of birth and date of assessment in hospital for SARS-CoV-2 infection.

#### Indices of multiple deprivation (IMD)

IMD scores were derived from postal codes for usual residence transcribed from hospital records. IMD quintile 1 represents the most deprived and quintile 5 the least deprived.

#### Critical care

Paediatric intensive care units (PICUs) are dedicated care settings providing the highest level of critical care for children and young people, who usually need invasive mechanical ventilation or support for two or more organ systems with a higher nurse to patient ratio. PICUs are usually located in regional tertiary centres or specialised hospitals. Paediatric high dependency units (HDUs) are for patients needing close monitoring and therapies for single organ system support, usually without invasive ventilation. HDUs are provided at tertiary hospitals and most district general hospitals.

#### Duplicates

Each child is represented once in the dataset. In cases where the child was readmitted, the admission with the highest level of care was retained. If readmissions required the same level of care, the earliest admission was retained. Where the child was transferred from one participating site to another during the same episode of care, their data were considered as one admission, retaining the first available vital signs and laboratory results and recording the highest level of treatments they had received.

#### Criteria for diagnosis of MIS-C

The case report form contained a Y/N variable for multisystem inflammatory syndrome in children (MIS-C) also known as paediatric inflammatory multisystem syndrome temporally associated with SARS-CoV-2 (PIMS-TS). We also adjusted the World Health Organisation (WHO) preliminary case definition for MIS-C, as previously described^2^ and searched free text in case report forms for “MIS-C”, “PIMS-TS” and “IVIg” (intravenous immunoglobulin). Patients identified using these three approaches were collated and sites contacted directly to clarify the diagnosis and to collect further details. Patients with pathogenic bacteria identified in blood or cerebrospinal fluid cultures and those with a diagnosis of appendicitis were censored from the MIS-C subgroup.

#### Incidental SARS-CoV-2

**Coded “yes”:**

- Patients where SARS-CoV-2 was noted to be incidental in free text.
- Patients with alternative reasons for admission, where free text notes no SARS-CoV-2 symptoms or asymptomatic.
- Patients where there is a clear primary reason for admission unrelated to any infection symptoms in free text e.g. overdose, road traffic accident.
- Patients where there is a clear primary reason for admission which relates to a specific focal infection not thought to be associated with SARS-CoV-2 e.g. septic arthritis, eczema herpeticum.

**Coded “no / unknown”:**

- No mention of other reason for admission in free text, no mention of asymptomatic or incidental finding
- Patient came to hospital for another reason (eg pre admission screen) but had SARS-CoV-2 symptoms and was admitted for this

Patients who had been coded “yes” for other reason for admission, but where it was not clear whether SARS-CoV-2 symptoms also contributed to presentation. For example:

- Appendicitis.
- Diabetic ketoacidosis.
- Systemic infection related admissions, where co-infection could be present.

#### Other reason for admission

**Coded “yes”**

Patients where free text includes reference to another acute condition which could have contributed to admission. This includes but is not limited to:

- Fractures, burns and other injuries.
- Eating disorder, self-harm, drug overdose, psychosis (note where free text states only depression or anxiety this was considered chronic, and not an alternative reason for admission unless stated otherwise).
- Surgical admissions including appendicitis.
- New presentation of type 1 diabetes.
- Neonatal jaundice.
- Patients admitted in labour or for elective caesarean section.

Patients where free text includes reference of admission related to a chronic condition. This includes, but is not limited to:

- Exacerbation of inflammatory bowel syndrome.
- Diabetic ketoacidosis in known diabetic.
- Elective admissions for diagnostic investigations.
- Elective admissions for surgical procedures or chemotherapy.

**Coded “no / unknown”**

Patients where free text includes reference to acute symptoms/presentation which could also represent symptoms of SARS-CoV-2. Examples:

- Febrile convulsions.
- Seizures in known epilepsy.
- Acute exacerbation of asthma.
- Gastroenteritis.

#### Missing data

Capacity to enrol into the current study was limited by staff availability, especially during admission surges research staff were redeployed to clinical activities. We did not impute missing data. All patients were admitted at least two weeks prior to the date of data extraction to minimise missing data. Denominators differ between analyses owing to incomplete data recorded for some variables. The research team undertook data cleaning and source verification to ensure the data extracted and analysed were as accurate as possible.

## Supplementary Results

### Assessment of potential sources of bias

In total, 764 CYP were admitted during the first wave (17^th^ January to 31st July 2020) and 1,280 during the second wave (1^st^ August 2020 to 31st January 2021) across a total of 187 sites of which 23 had access to an onsite paediatric intensive care unit (PICU) (*Supplementary Figure C* and *Supplementary Table A*). As reporting to ISARIC is not mandatory, we examined whether sites with onsite PICUs reported more CYP in one wave than another, which might bias the severity of illness in reported patients. 118 hospitals reported paediatric patients to ISARIC in both the first and second waves, 34 more reported in the first wave only and 35 in the second wave only. The proportion of patients reported from hospitals with access to an onsite PICU did not differ between the waves (37.0% (283/764) in the first wave vs 35.5% (455/1280, p = 0.53) in the second wave).

We then compared the number of CYP reported to ISARIC against the numbers of local SARS-CoV-2 cases identified by Pillar 1 and 2 testing by Public Health England across NHS regions (*Supplementary Figure D*). In the ISARIC dataset, regional peaks were seen in the Midlands in November 2020 and in London in December 2020 which closely mirrored those reported by Public Health England at the same time points, indicating that ISARIC also captured localised SARS-CoV-2 peaks across the UK.^3^

## Supplementary Tables

**Supplementary Table A.** Comparison of the number of sites reporting to ISARIC in the first and second waves against the number of patients reporting and whether the hospital had an on-site PICU.

| Wave | Hospitals | Patients | Onsite PICU |
| --- | --- | --- | --- |
| First only | 34 | 99 | 2 |
| First and Second | 118 | 1747 | 20 |
| Second only | 35 | 198 | 1 |

**Supplementary Table B.**
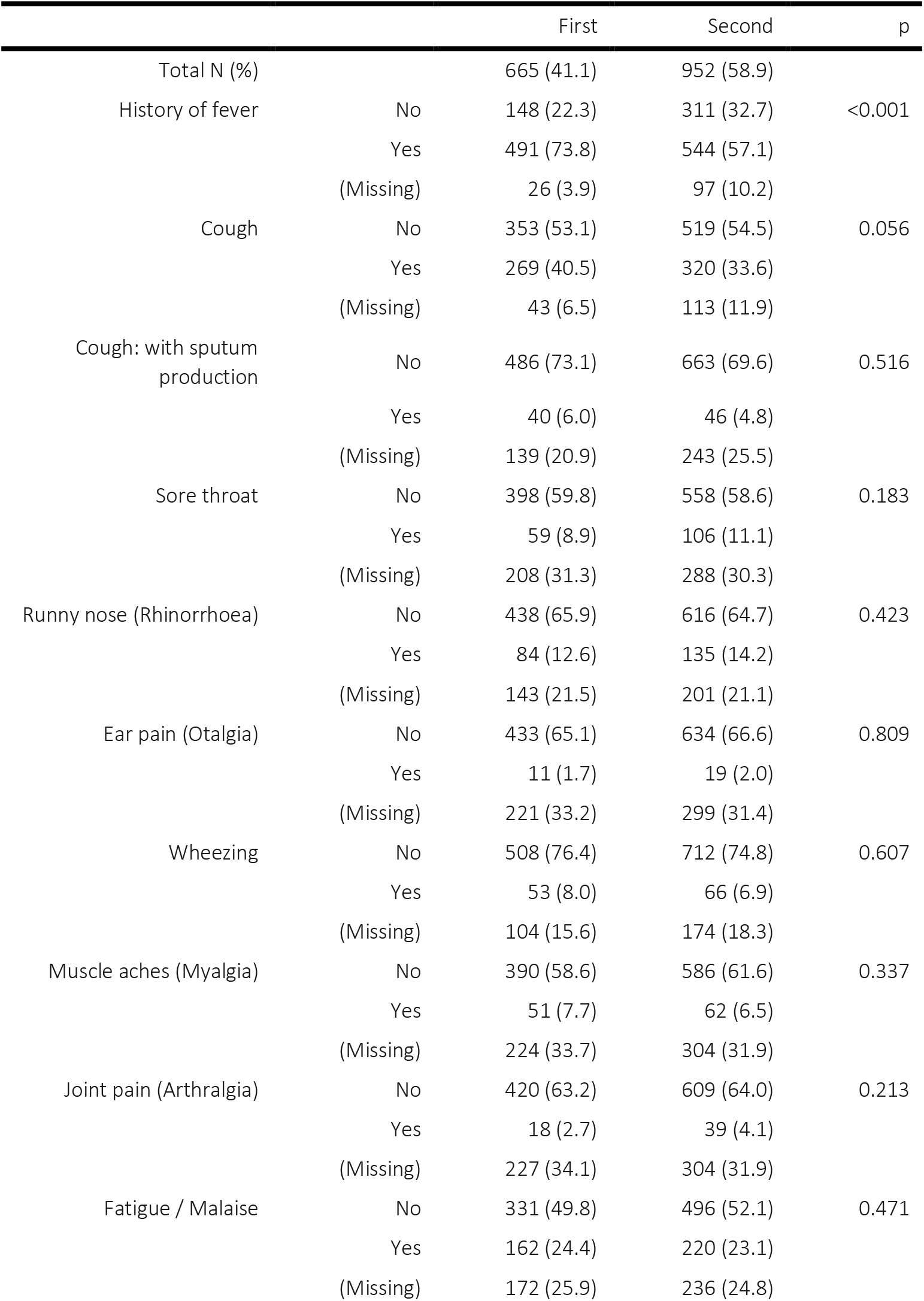
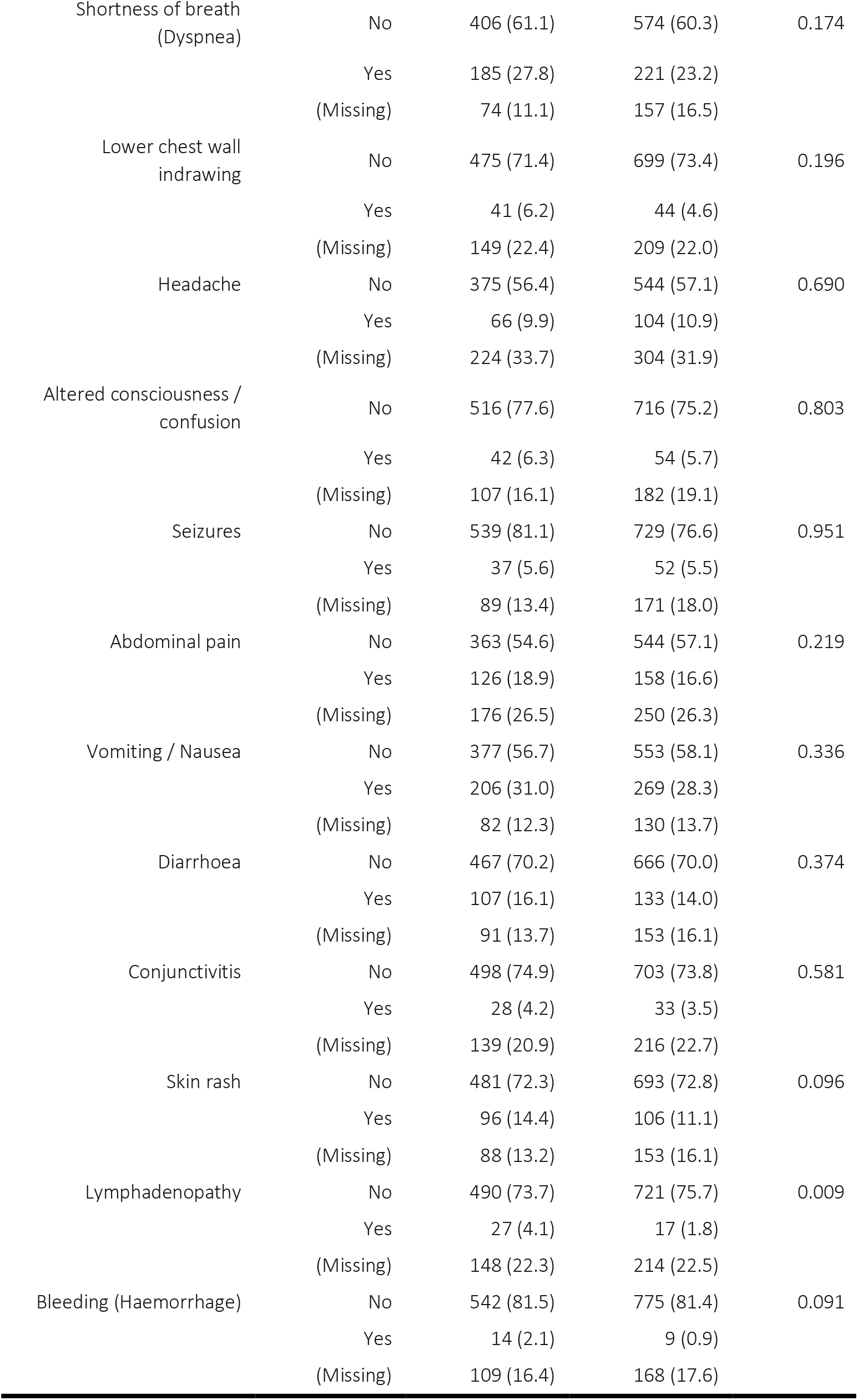
Presenting symptoms by wave, with CYP with asymptomatic / incidental SARS-CoV-2 infections excluded.

**Supplementary Table C.**
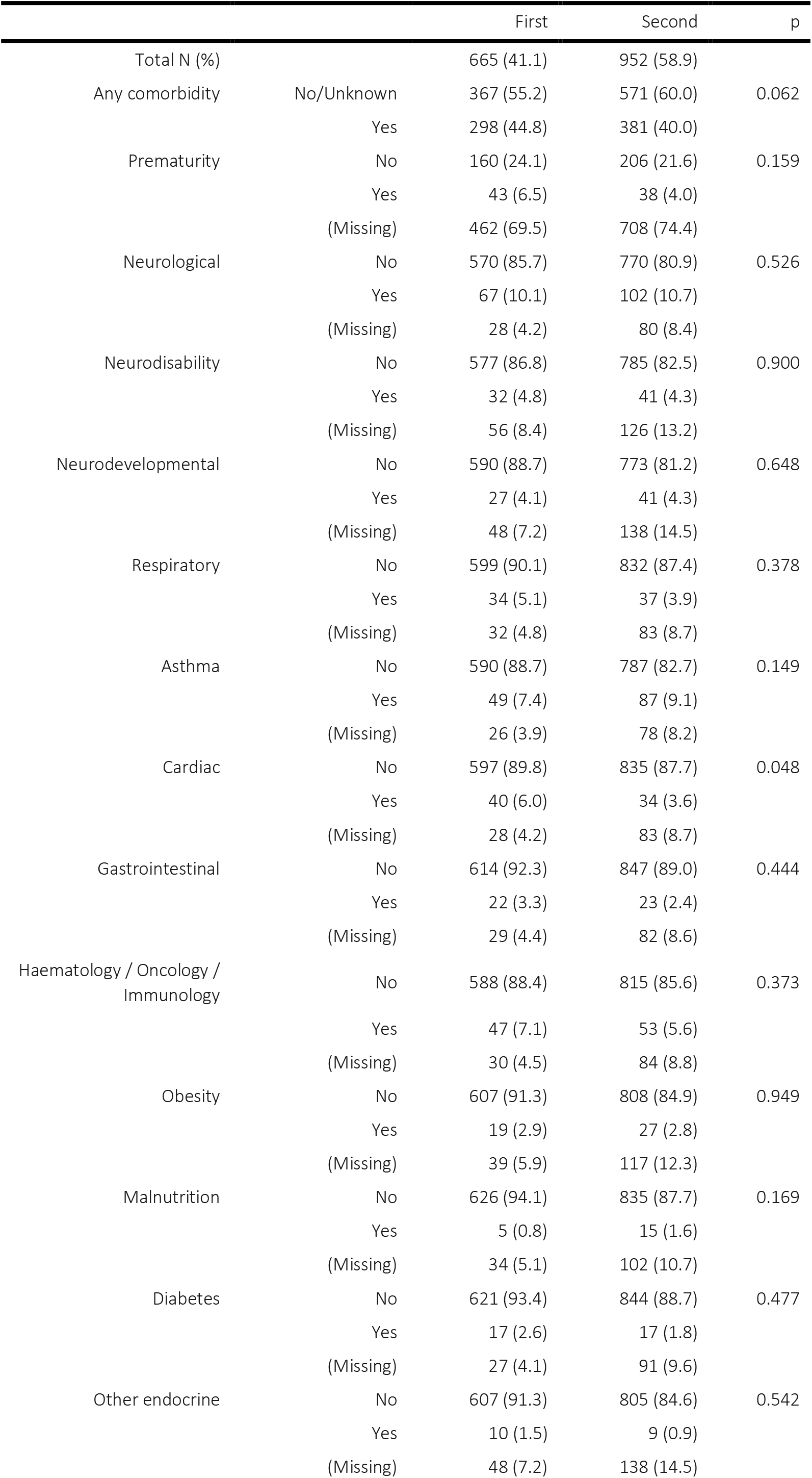
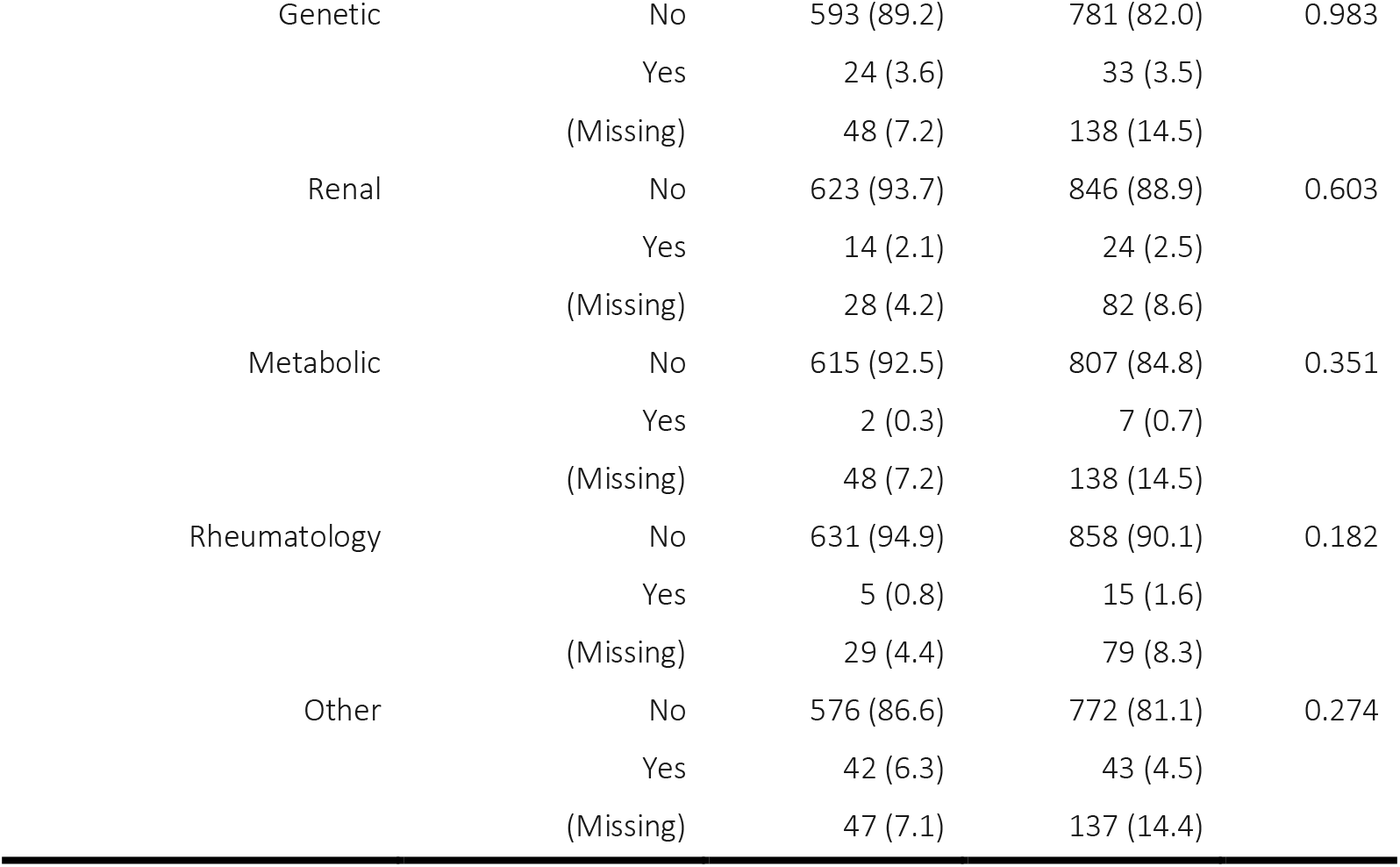
Comparison of comorbidities across the two waves, CYP with asymptomatic or incidental SARS-CoV-2 excluded.

**Supplementary Table D.**
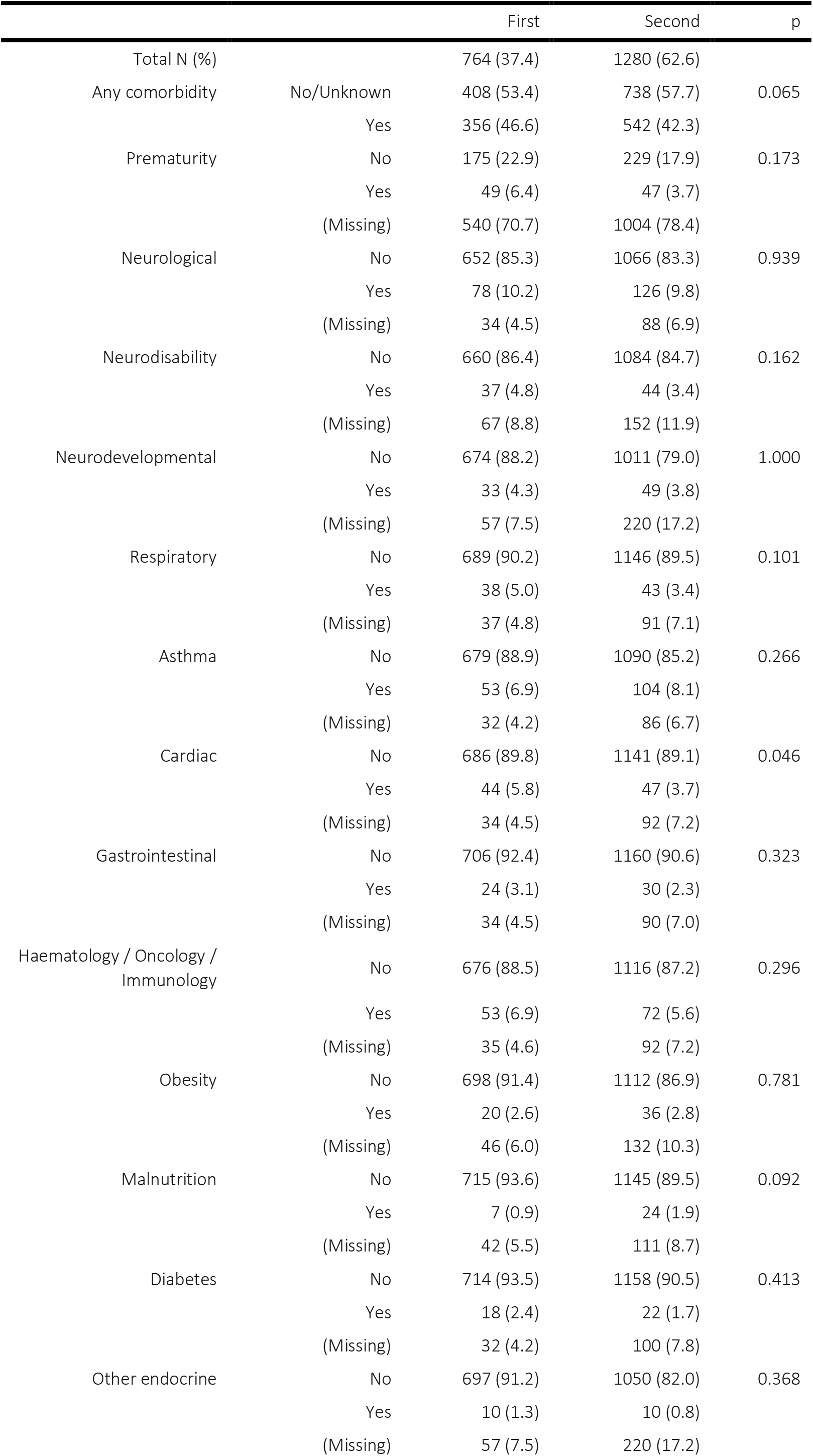
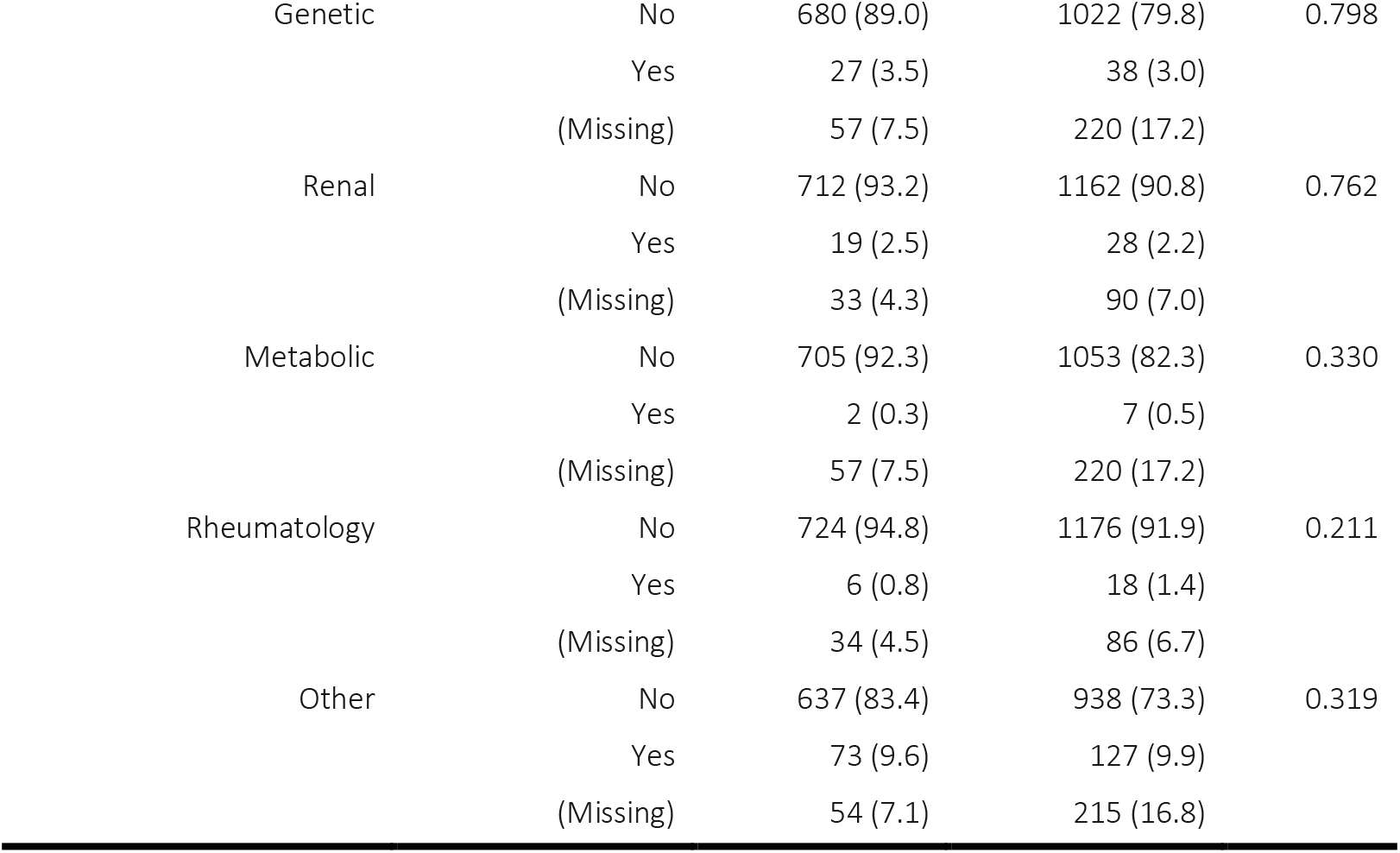
Comparison of comorbidities for the whole cohort (i.e. <u>includes</u> asymptomatic or incidental SARS-CoV-2) across the two waves.

**Supplementary Table F.**
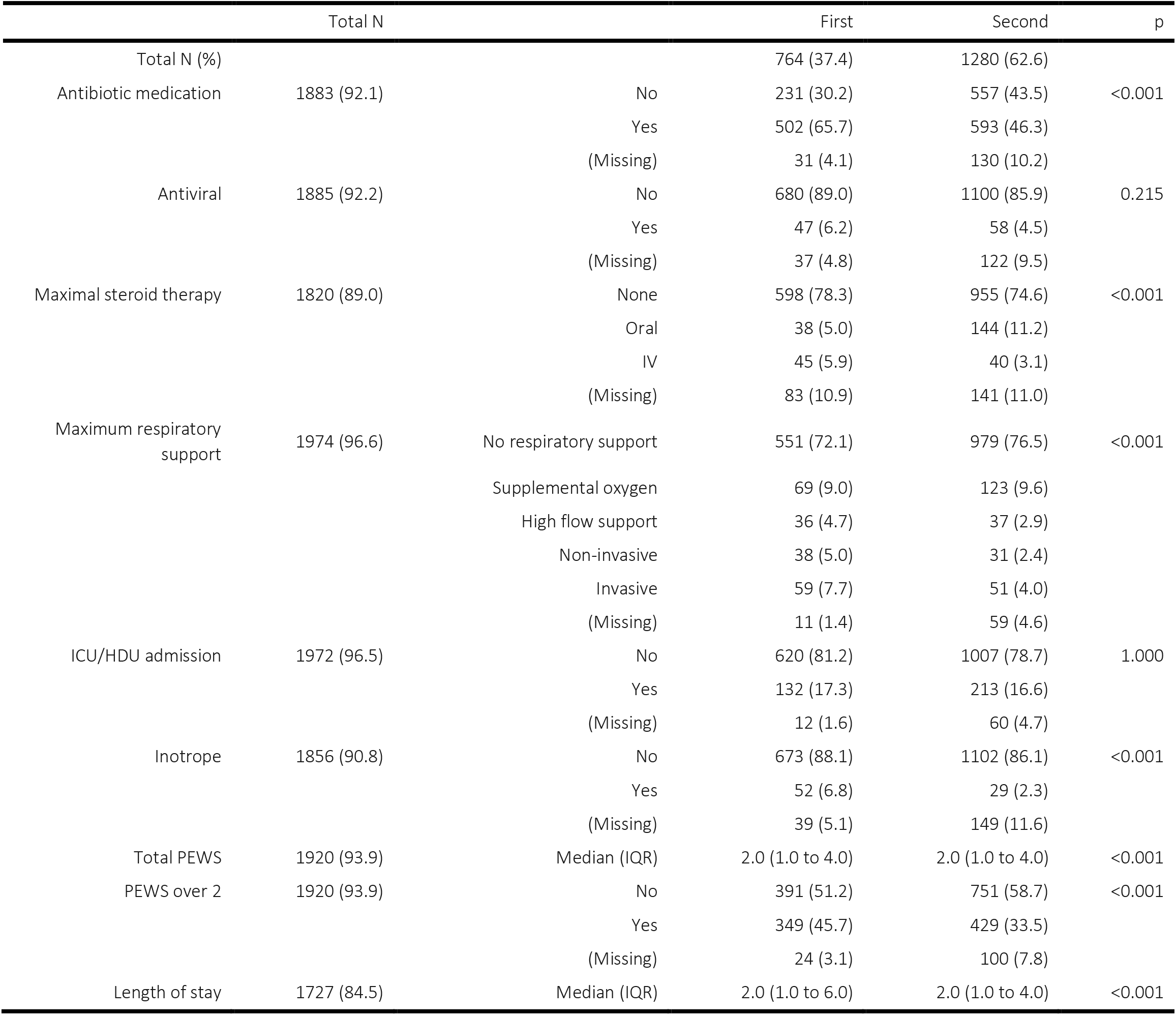
Comparison of treatments received by children by wave across the whole cohort (i.e. <u>includes</u> asymptomatic and incidental SARS-CoV-2). ICU = intensive care unit, HDU = high dependency unit. PEWS = Paediatric Early Warning Score at presentation.

**Supplementary Table G.**
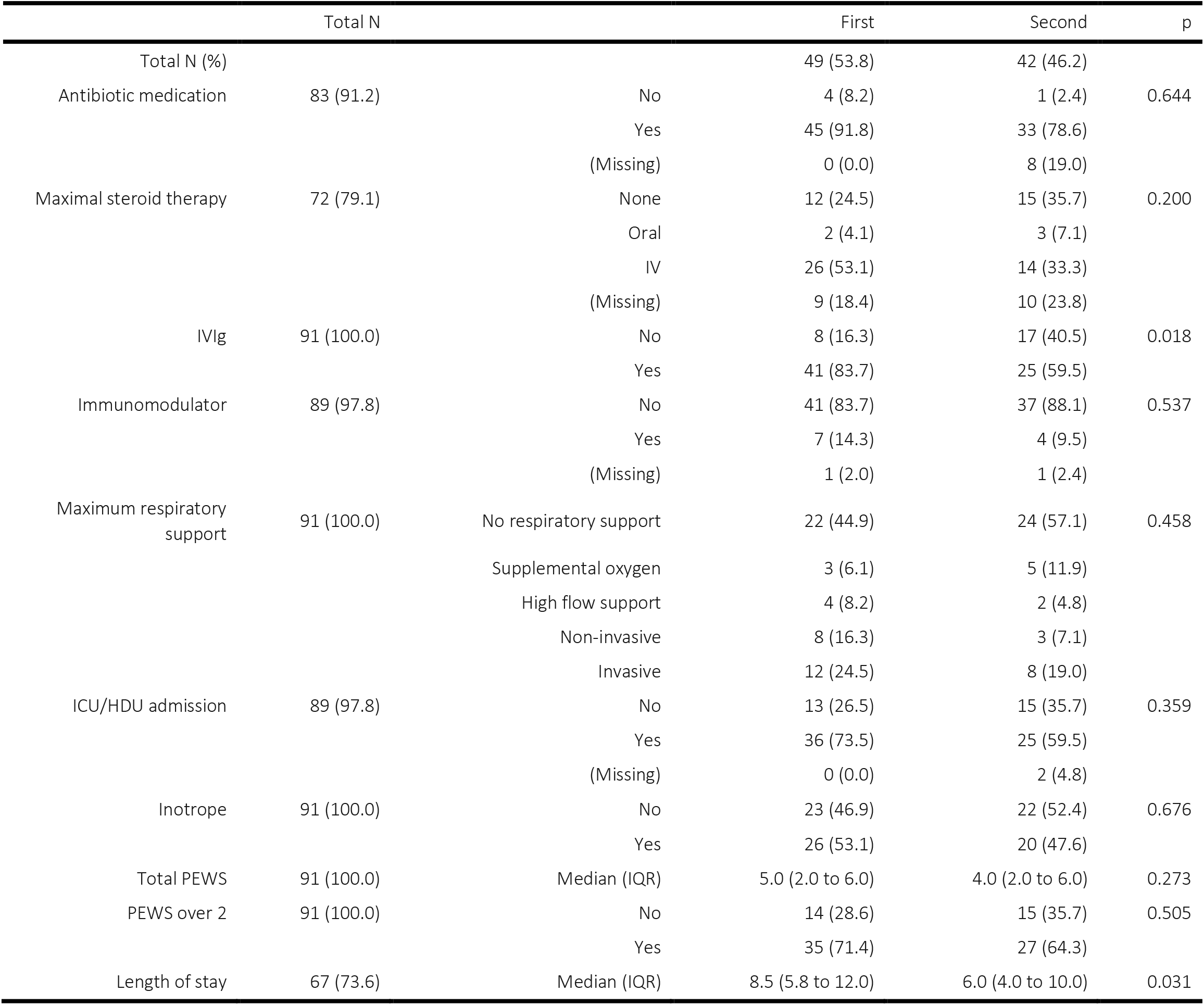
Treatments received by children with MIS-C by wave. ICU = intensive care unit, HDU = high dependency unit, PEWS = Paediatric Early Warning Score at presentation, IQR = interquartile range.

**Supplementary Table H.**
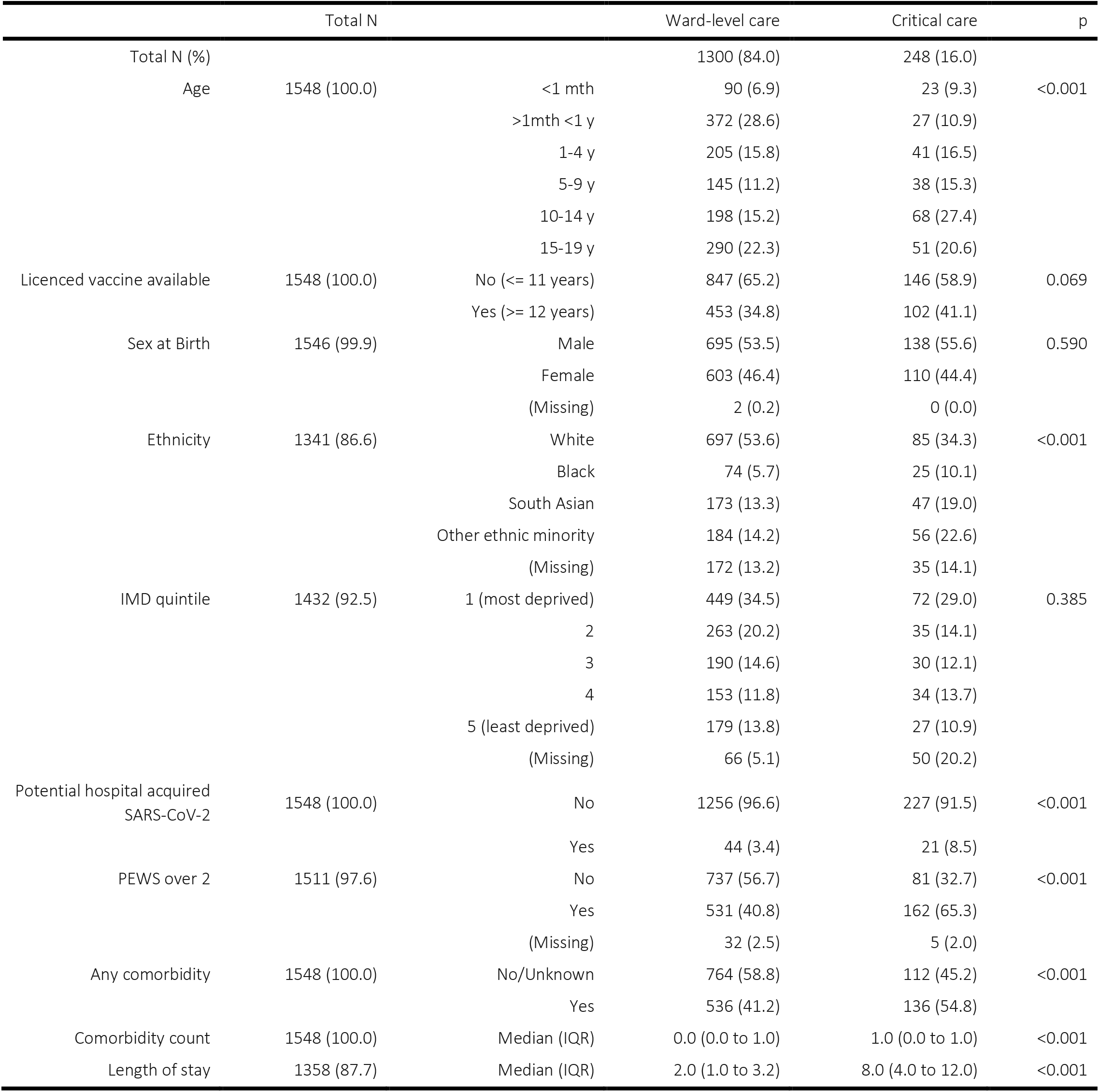
Demographics and key clinical characteristics of children stratified by critical care admission (excluding asymptomatic or incidental SARS-CoV-2 infections but <u>including</u> those with MIS-C). IMD = indices of multiple deprivation. PEWS = Paediatric Early Warning Score at presentation.

**Supplementary Table I.**
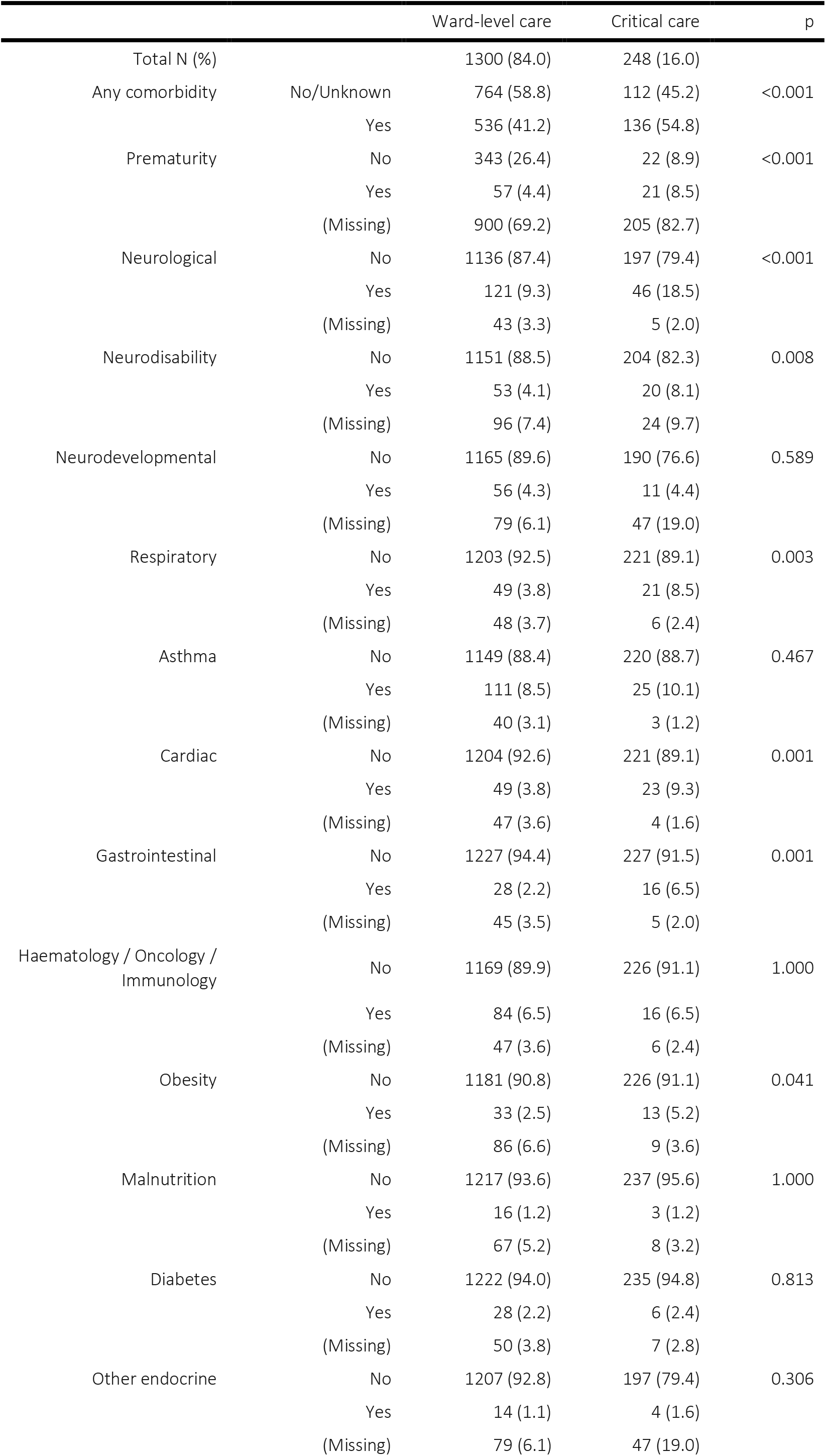
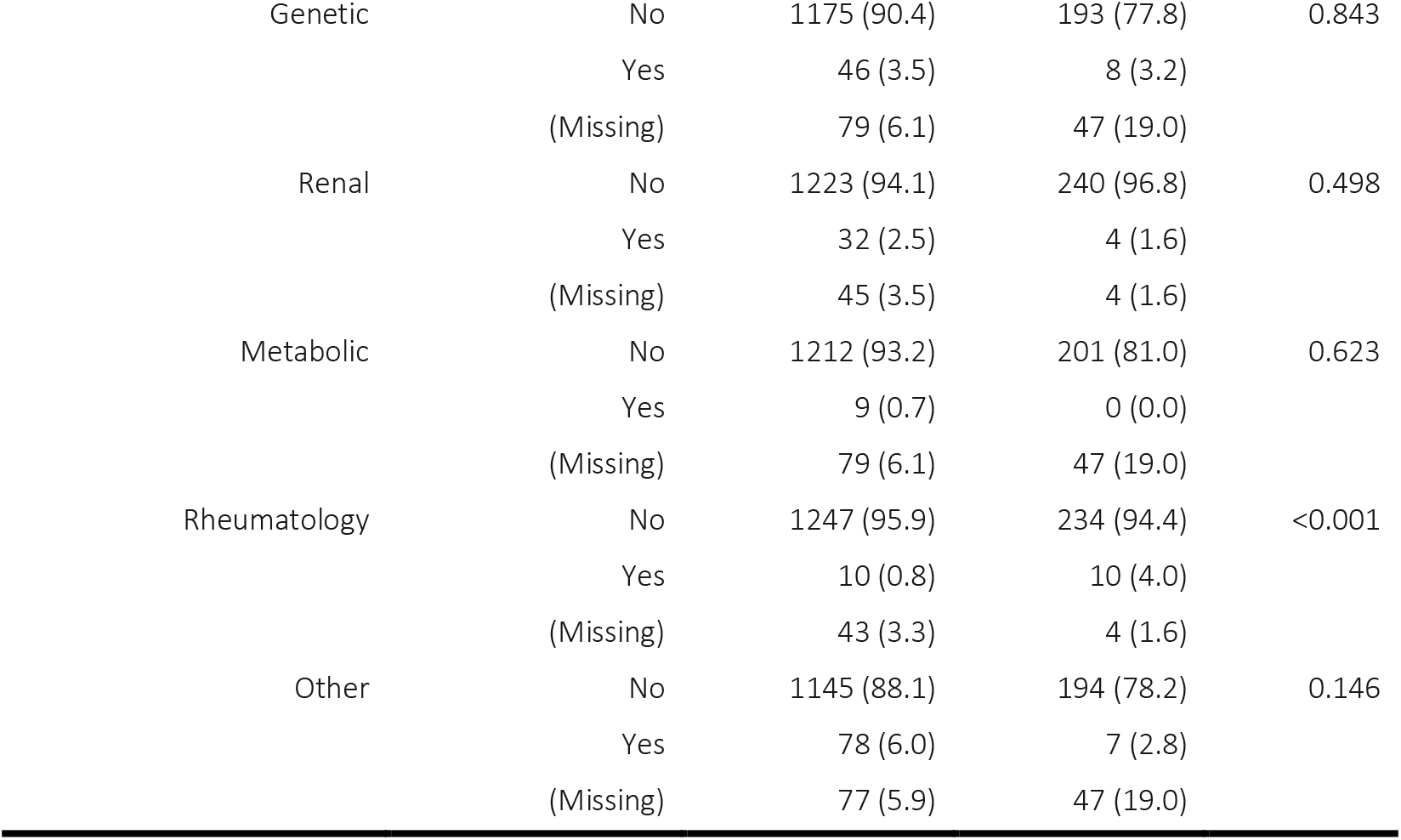
Comorbidities of CYP stratified by critical care admission excluding asymptomatic or incidental SARS-CoV-2 infections but <u>including</u> those with MIS-C.

**Supplementary Table J.**
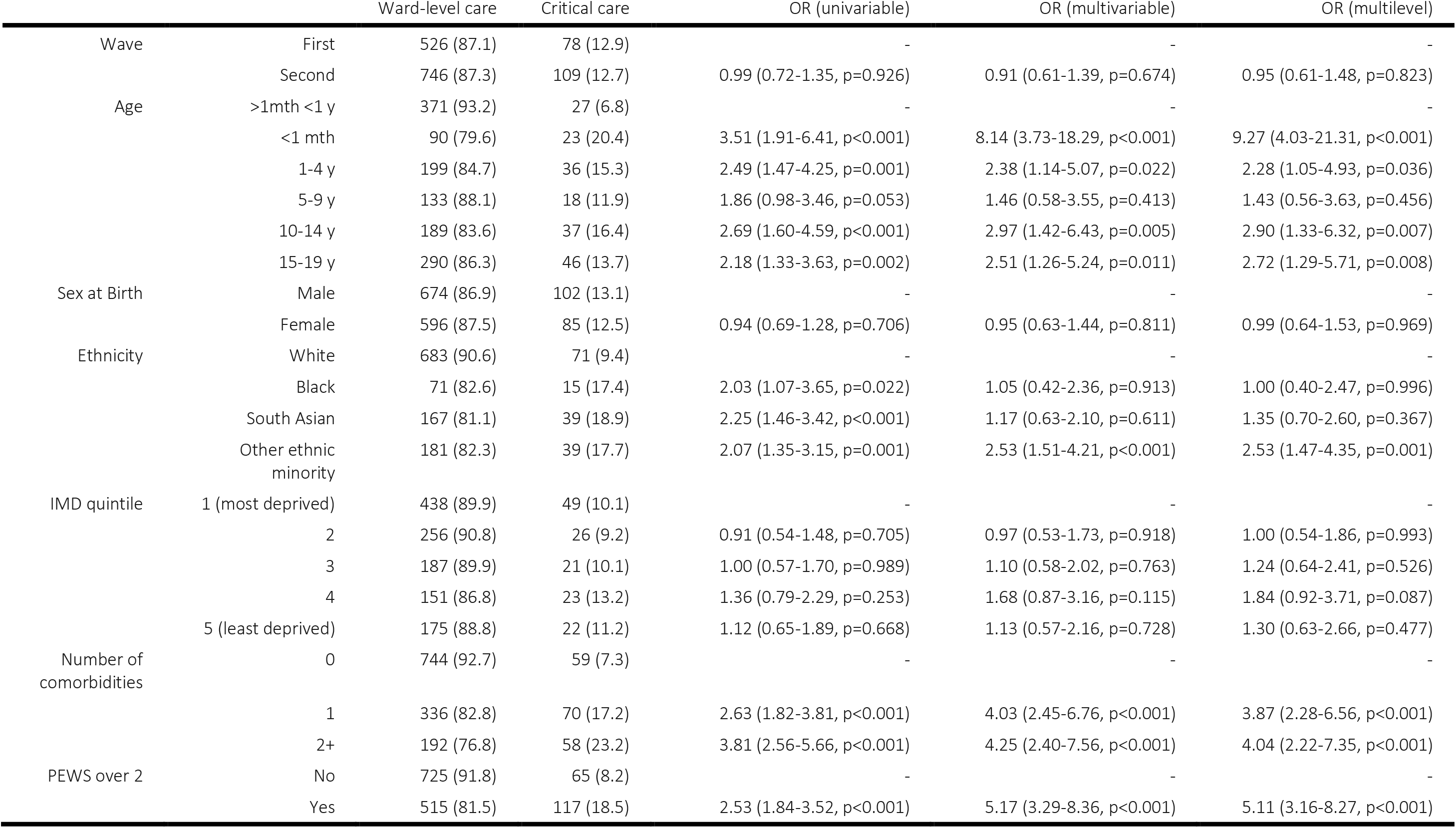
Multivariable analysis of factors associated with admission to critical care unit (excluding asymptomatic and incidental SARS-CoV-2 infections, and patients with Multisystem inflammatory syndrome in children (MIS-C)). Row percentages. IMD = Index of multiple deprivation. PEWS = Paediatric Early Warning Score at presentation. Odds ratios (OR) and 95% confidence intervals are presented.

**Supplementary Table K.**
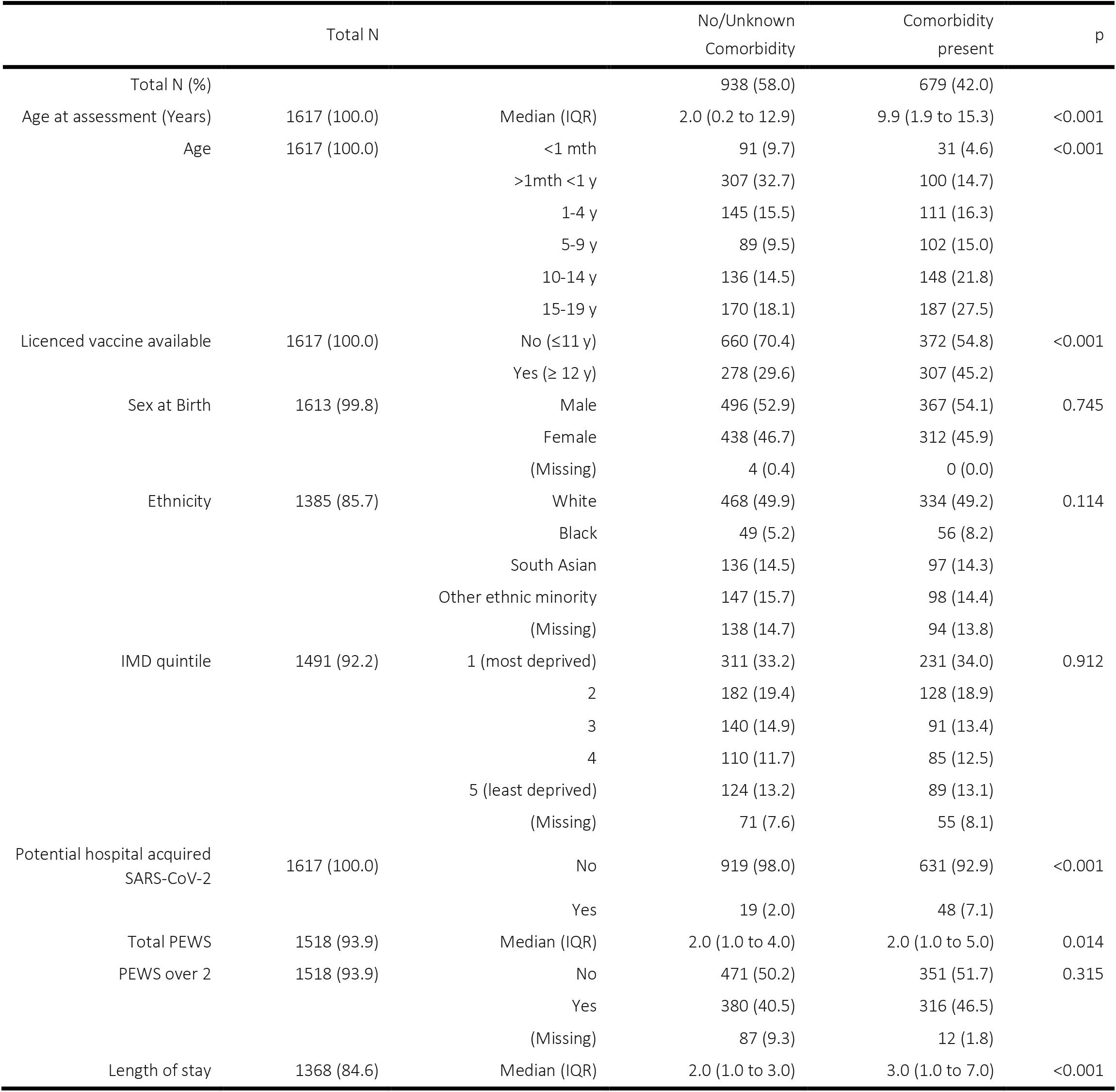
Demographics and key clinical characteristics of CYP stratified by comorbidity (patients with asymptomatic or incidental SARS-CoV-2 infections excluded). Column percentages by sub-group. IMD = Indices of multiple deprivation. PEWS= Paediatric Early Warning Score at presentation. IQR = Interquartile range

**Supplementary Table L.**
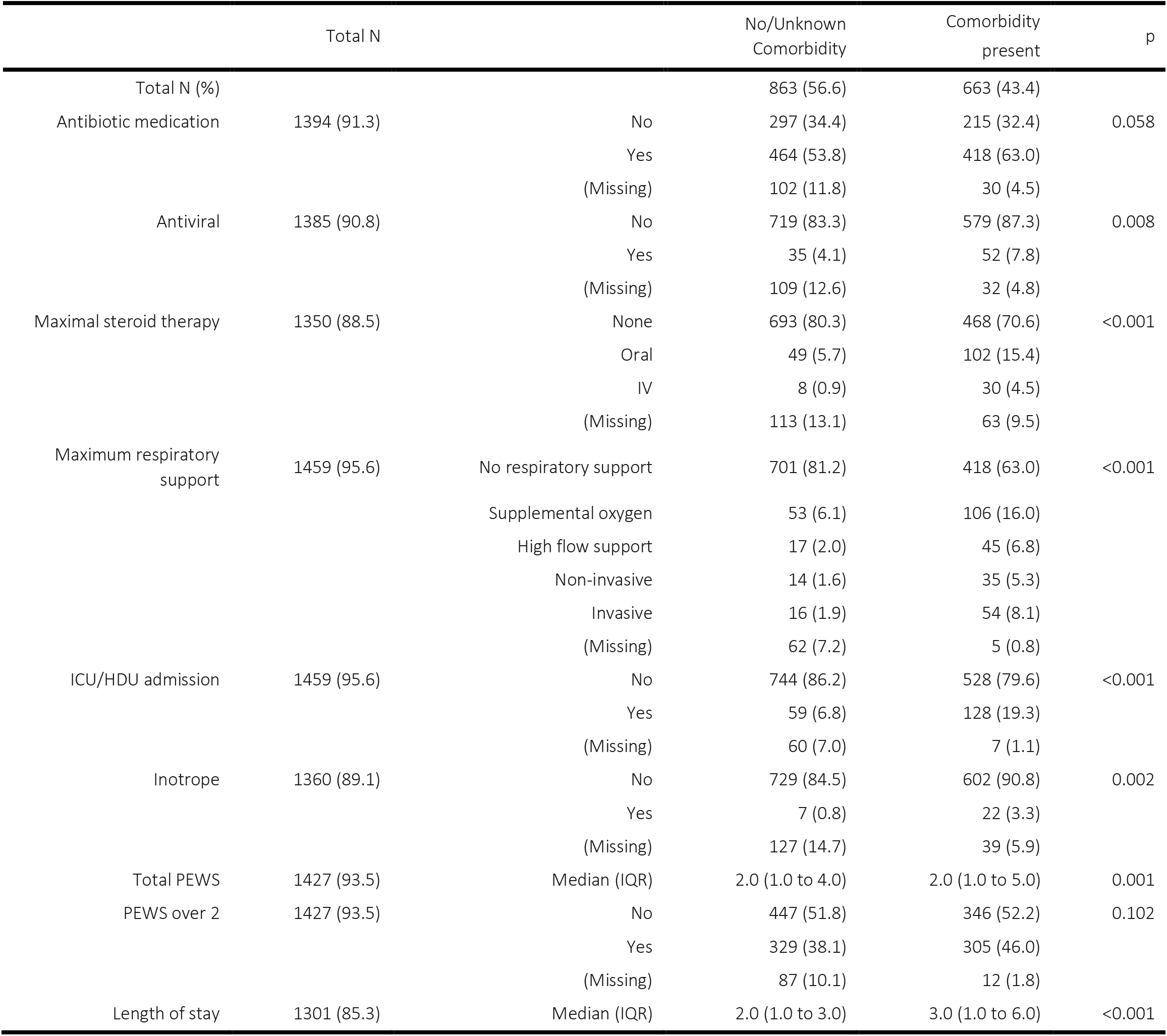
Treatments received stratified by comorbidity (excluding patients with asymptomatic or incidental SARS-CoV-2 infections and those with Multisystem Inflammatory Syndrome in Children (MIS-C)). ICU = intensive care unit. HDU = high dependency unit. PEWS= Paediatric Early Warning Score at presentation.

**Supplementary Table M.**
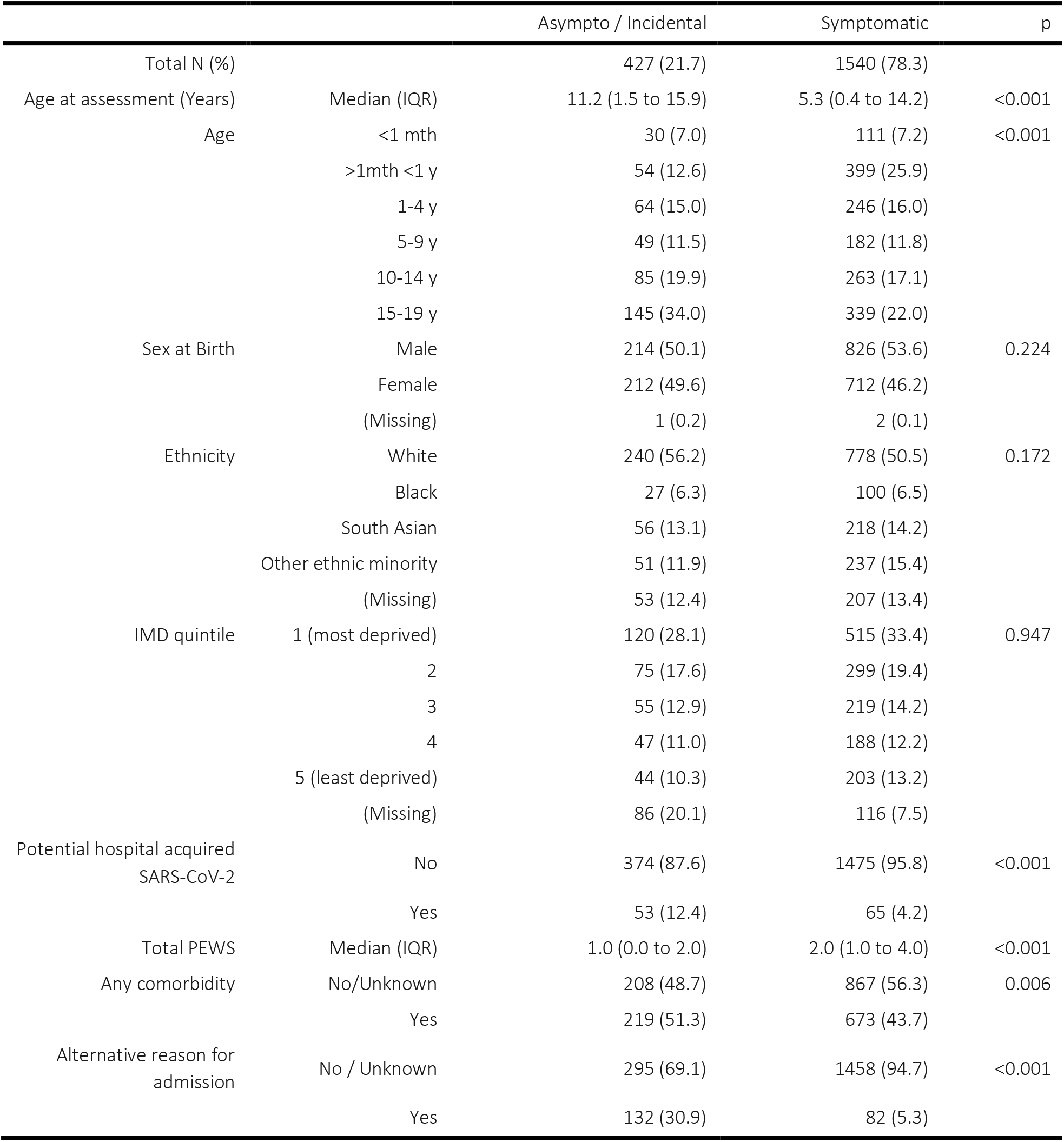
Demographics and key clinical characteristics of children with asymptomatic or incidental SARS-CoV-2 infections compared to those who were symptomatic. Column percentages by subgroup. IMD = Indices of multiple deprivation. PEWS= Paediatric Early Warning Score at presentation. IQR = Interquartile range

## Supplementary Figures

**Supplementary Figure A.**
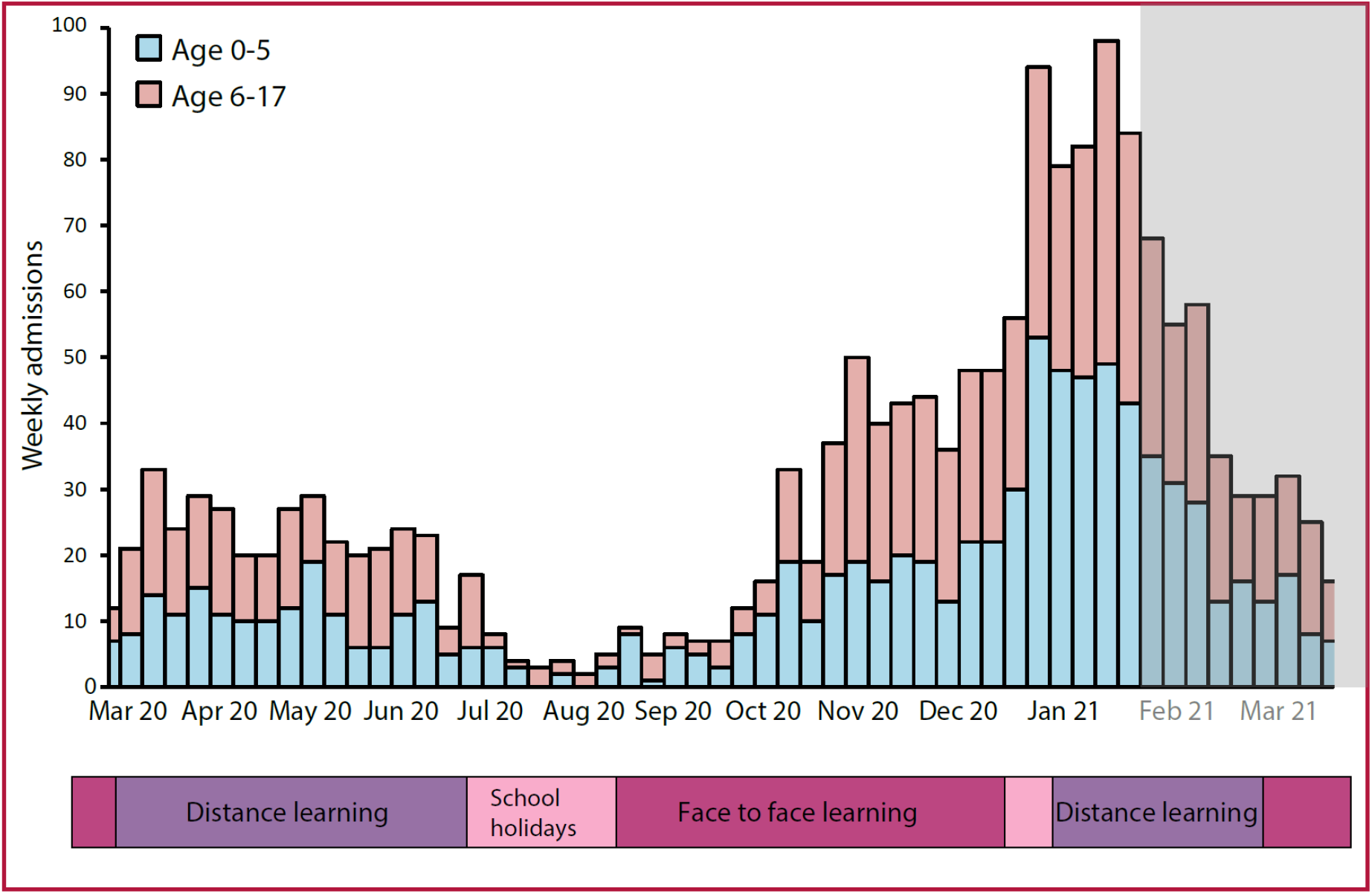
Timeline of major lockdown events in England against SARS-CoV-2 admissions for patients under 18 years across England (admissions data from NHS England).^4^ Magenta = face to face learning, purple = distance learning, pink = school holidays. This ISARIC4C analysis spans the period indicated by a white background. (NB. ISARIC-4C also includes data from Scotland and Wales)

**Supplementary Figure B.**
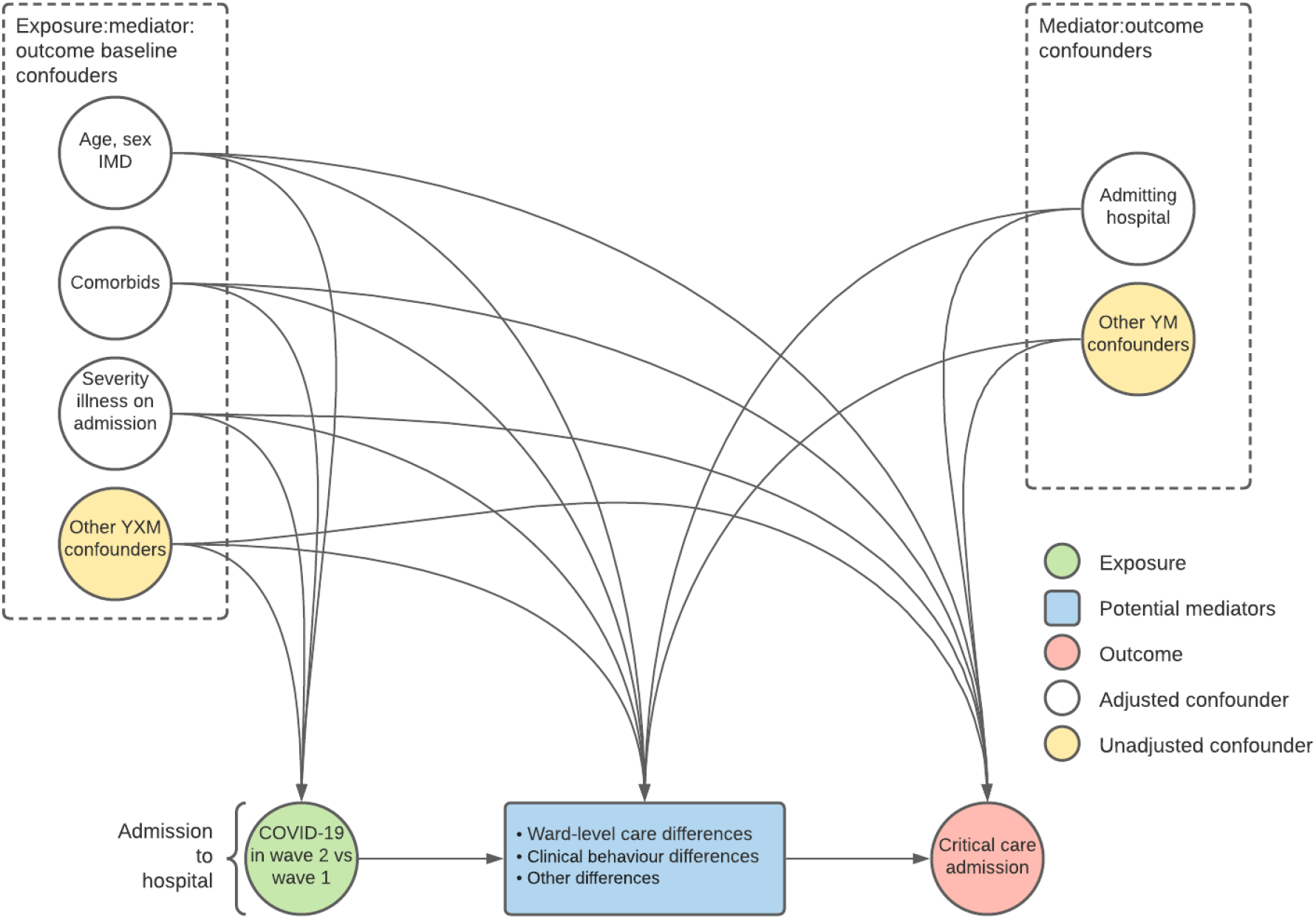
Directed acyclic graph of factors associated with critical care admission for construction of multivariable analysis

**Supplementary Figure C.**
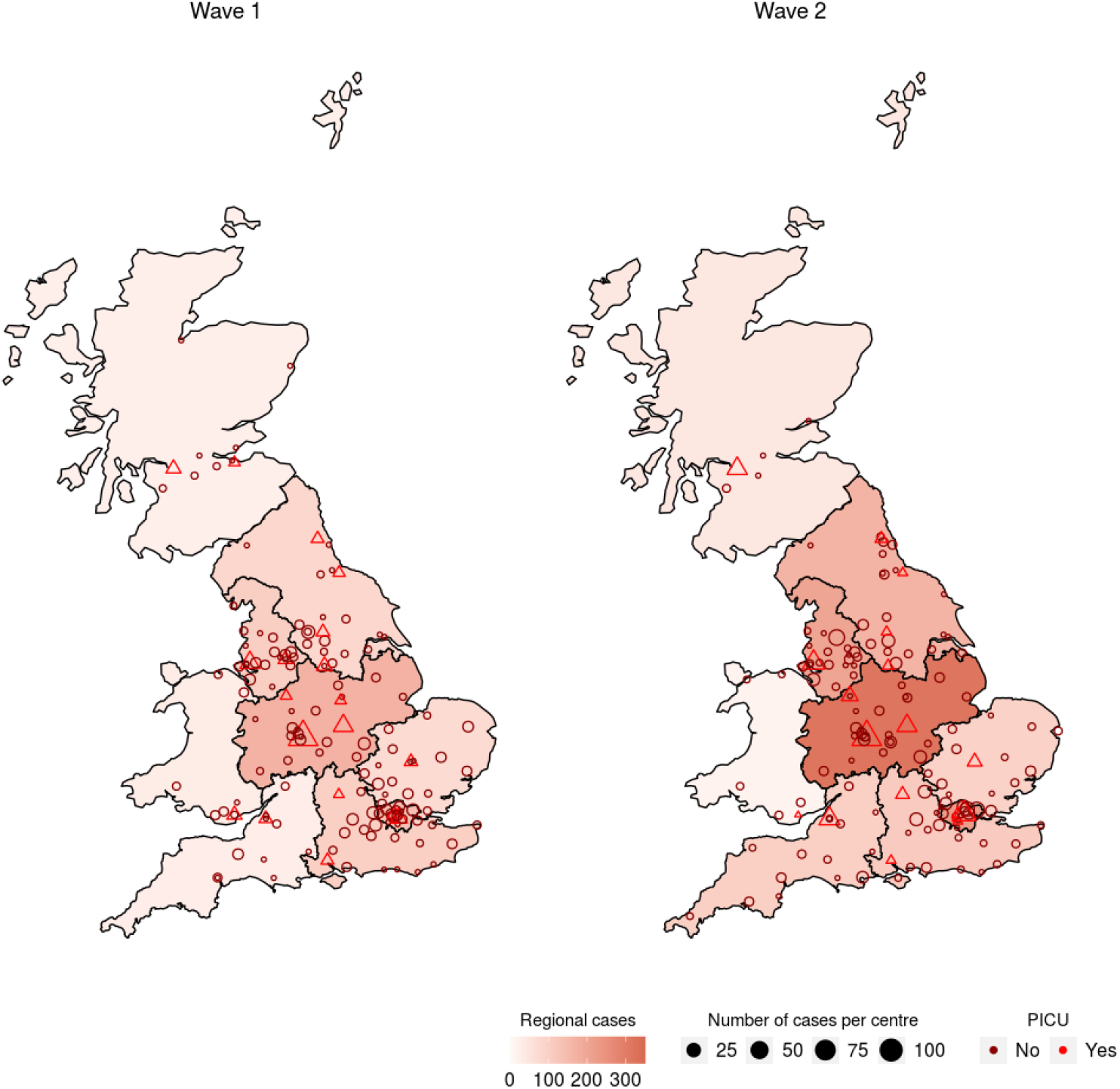
Map of sites of patient enrolment and cases by site. Sites with access to an onsite PICU are represented with triangles and those without as circles.

**Supplementary Figure D.**
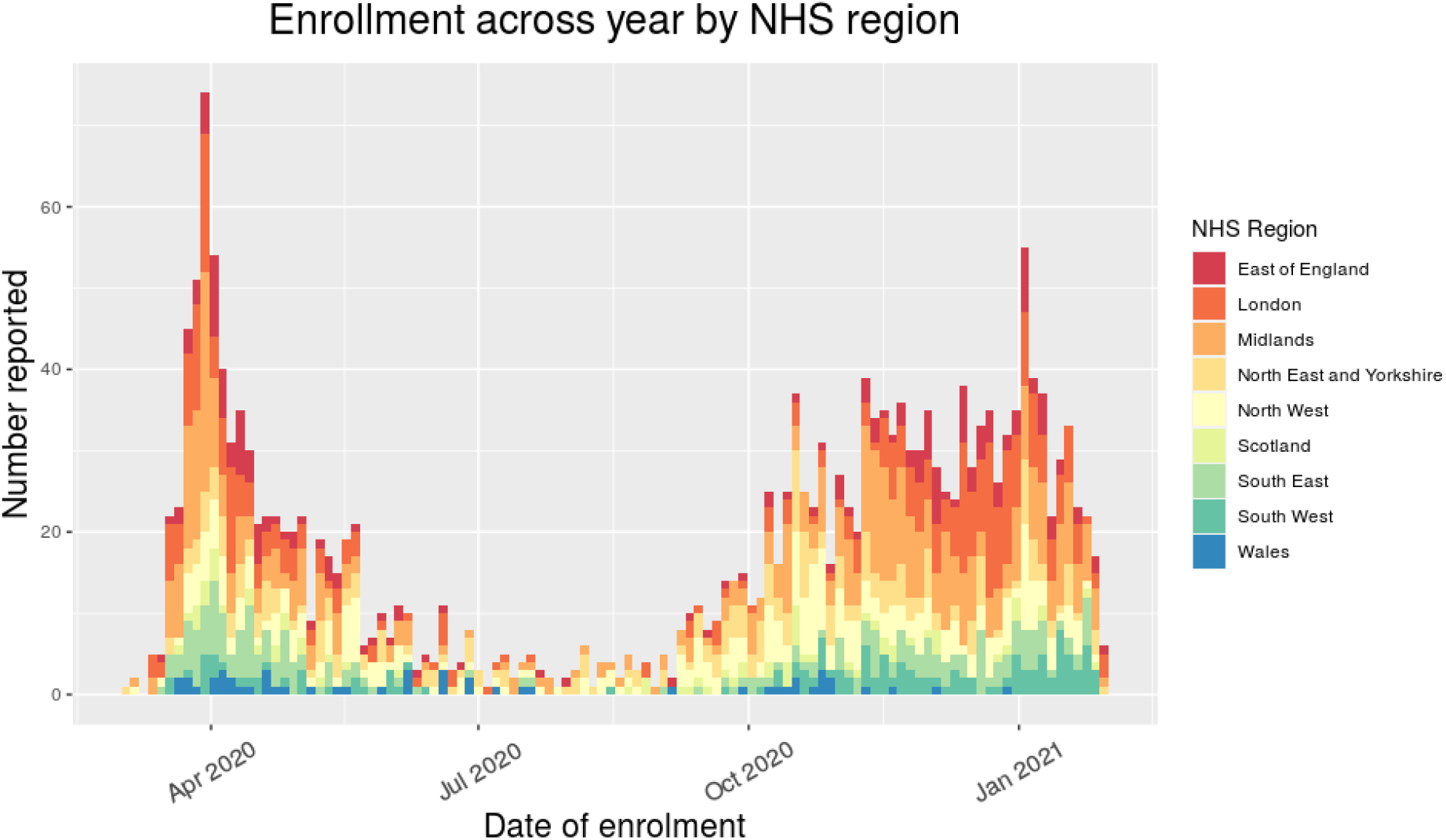
Reporting of patients <19 years to ISARIC4C by NHS region. Regional peaks can be seen in November in the Midland (orange) and London in December (dark orange). These peaks closely mirrored those reported by Public Health England at the same time points.^3^

**Supplementary Figure E.**
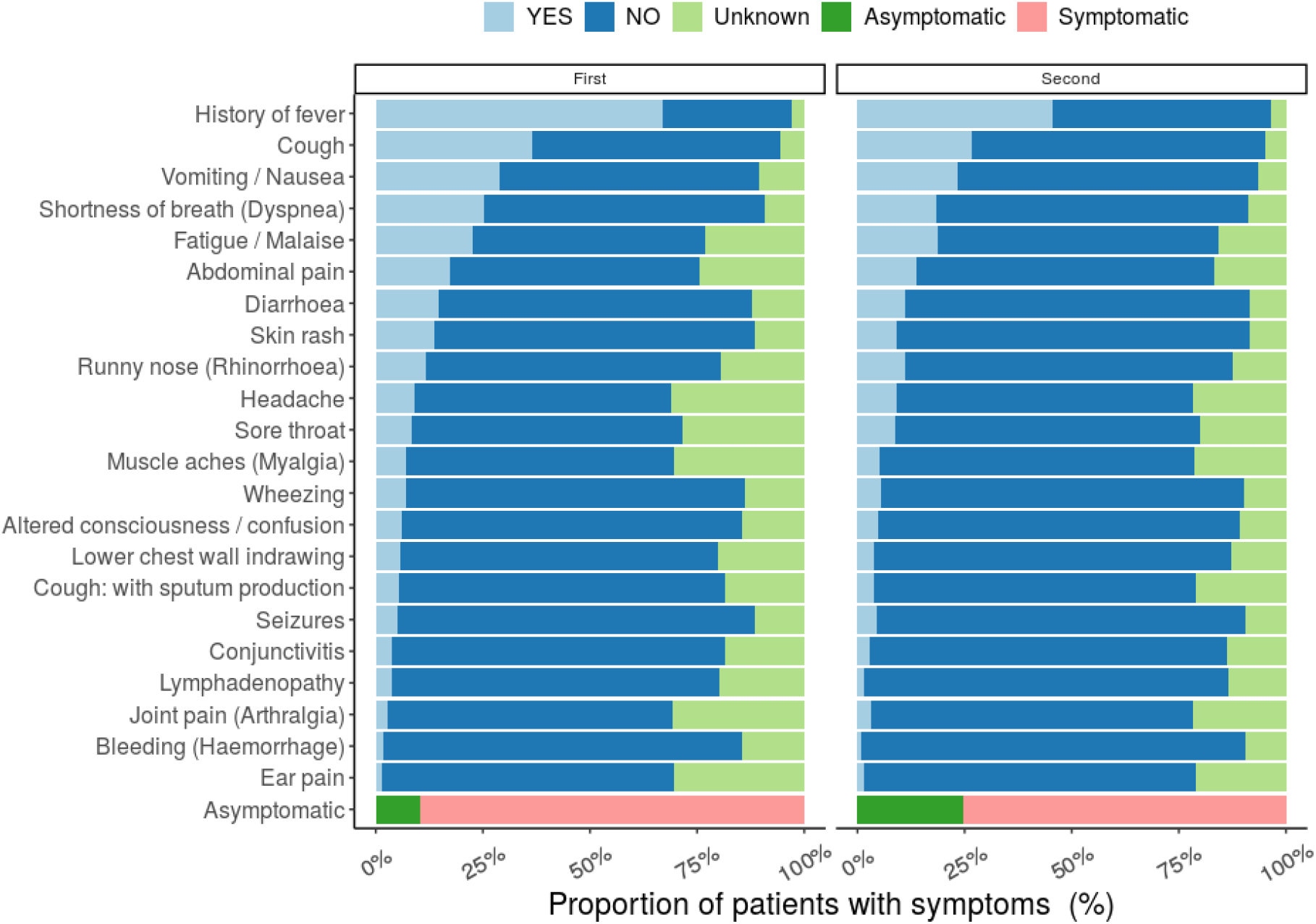
Comparison of symptoms at presentation (whole cohort analysis).

**Supplementary Figure F.**
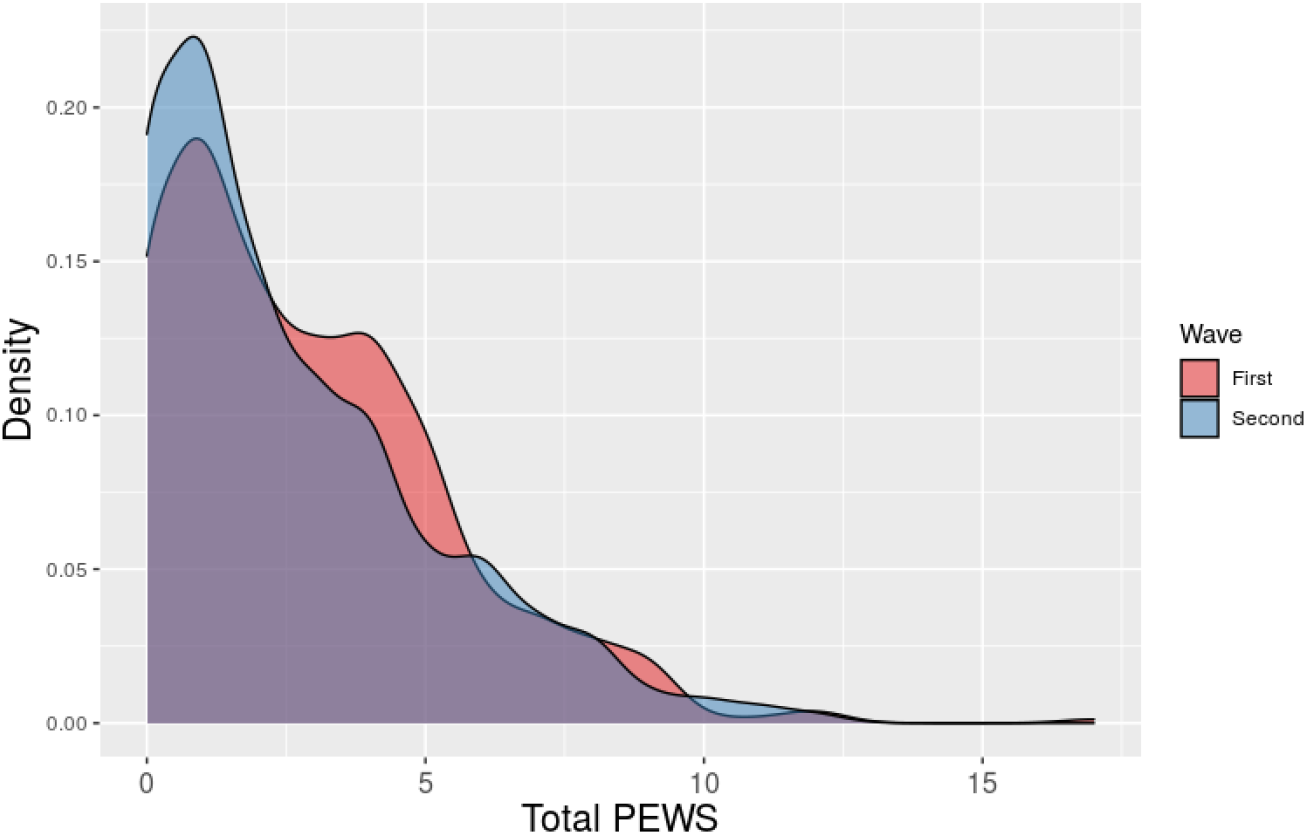
Paediatric Early Warning Score (PEWS) at presentation compared across the two waves (asymptomatic or incidental SARS-CoV-2 infections and patients with MIS-C excluded).

**Supplementary Figure G.**
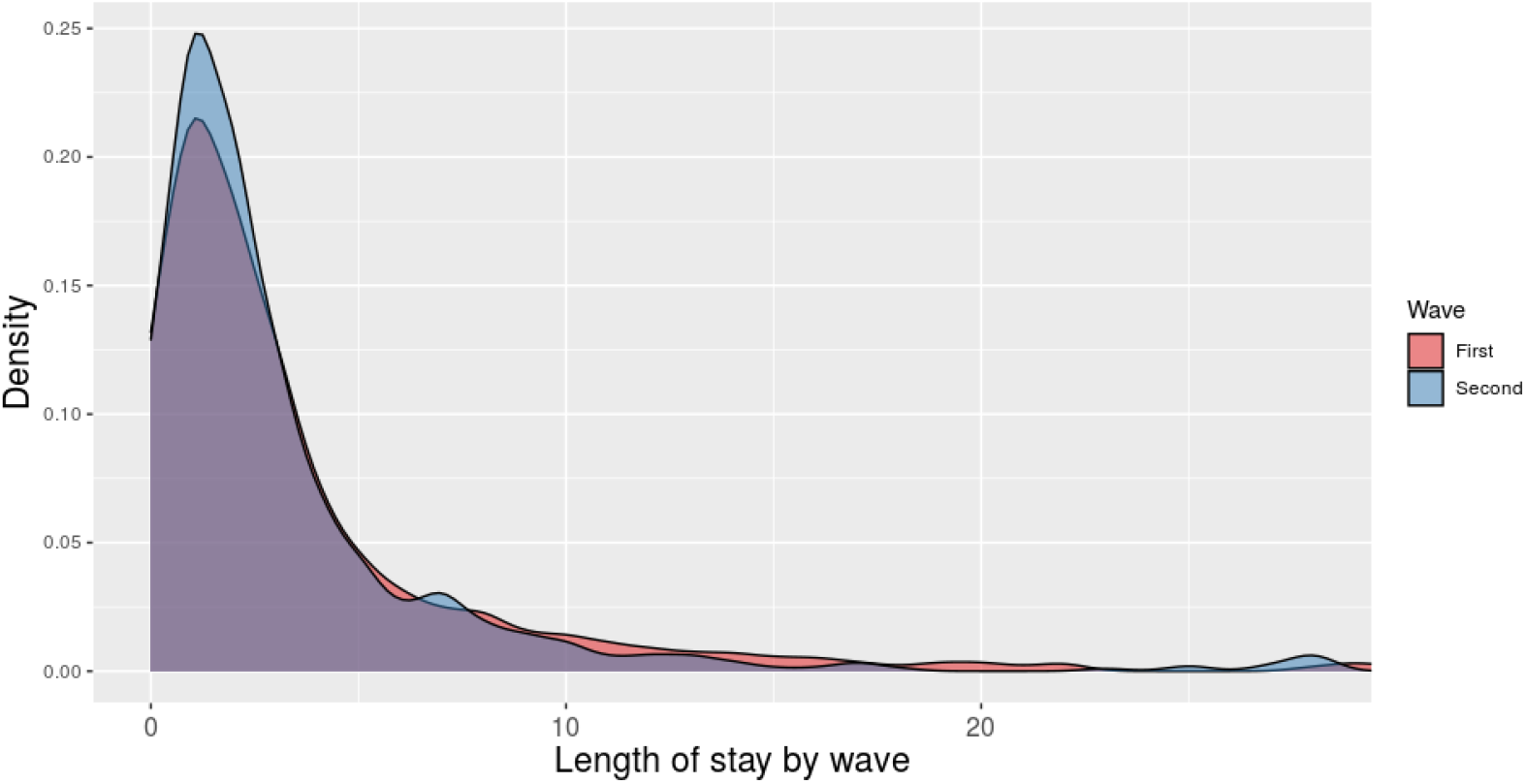
Length of stay compared across the two waves (asymptomatic / incidental SARS-CoV-2 infections and patients with MIS-C excluded)

**Supplementary Figure H.**
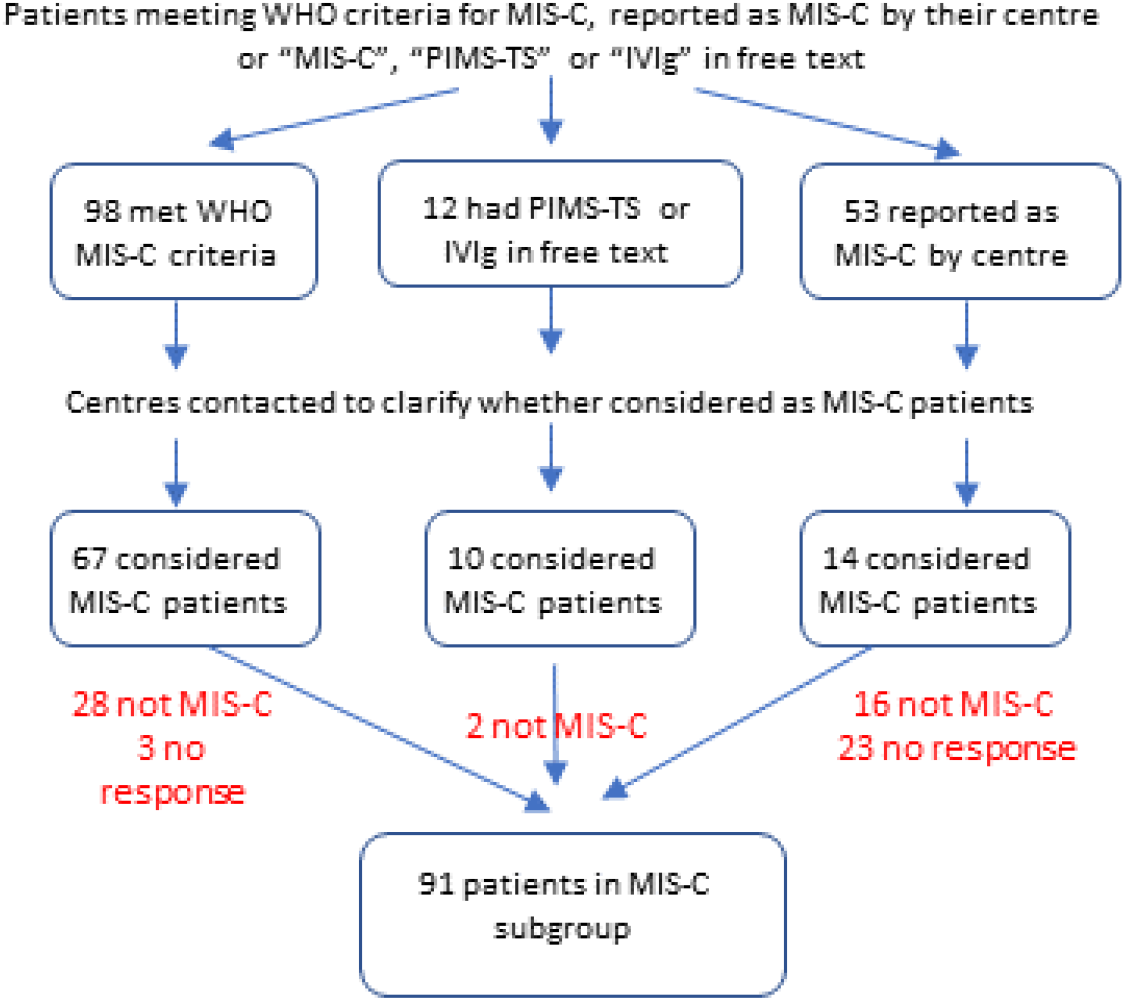
Flowchart for identification of patients with multisystem inflammatory syndrome (MIS-C). WHO = World Health Organisation, PIMS-TS = Paediatric Multisystem Inflammatory Syndrome – Temporally Associated with SARS-CoV-2, IVIg = Intravenous immunoglobulin.

**Supplementary Figure I.**
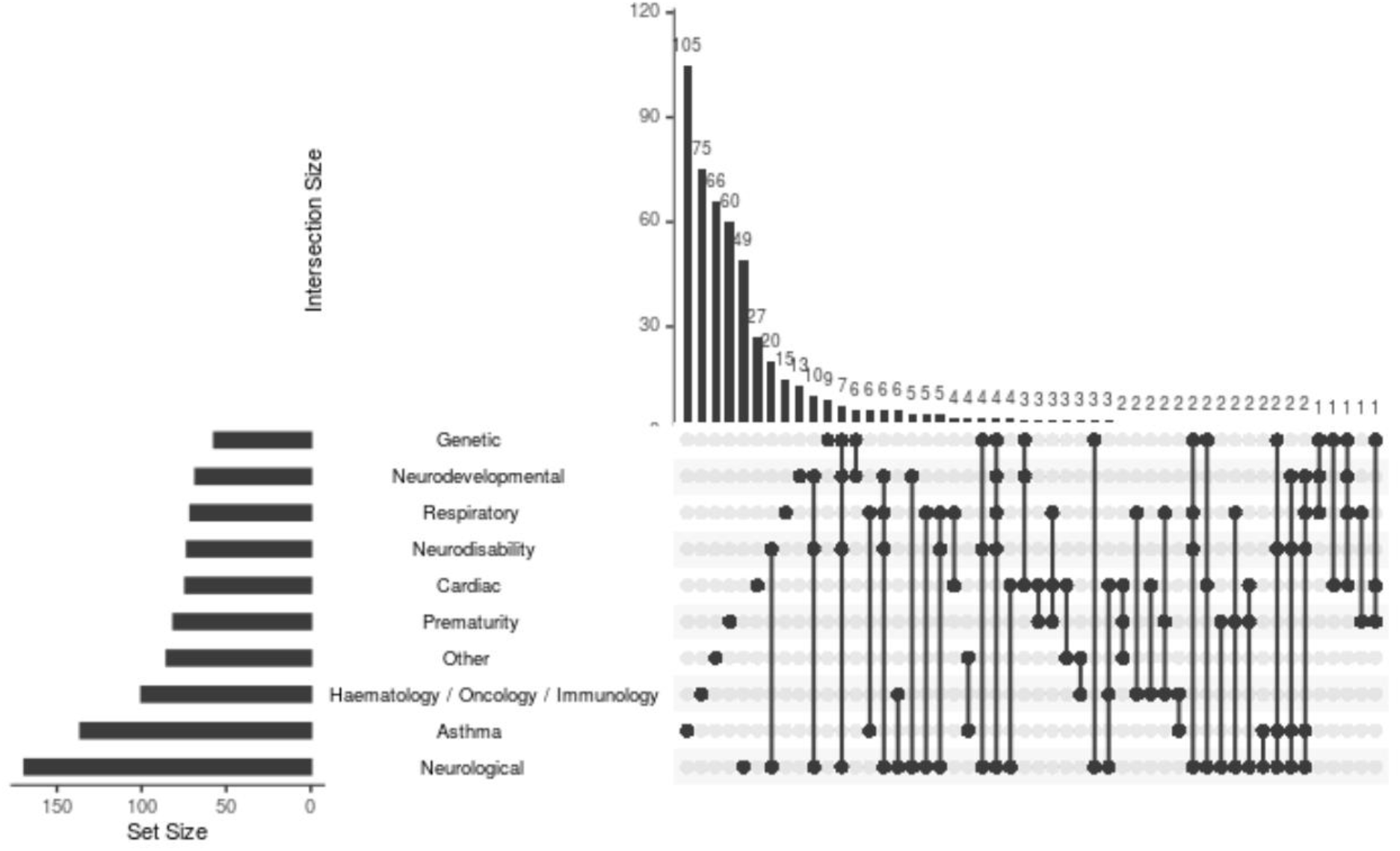
UpSet plot of patients comorbidities in patients admitted to critical care (excluding asymptomatic and incidental SARS-CoV-2 infections). Plot represents a visualisation of set intersections in the data.

## References

1. Ladhani SN, Amin-Chowdhury Z, Davies HG, et al. COVID-19 in children: analysis of the first pandemic peak in England. Arch Dis Child 2020; 105(12): 1180–5.

2. Swann OV, Holden KA, Turtle L, et al. Clinical characteristics of children and young people admitted to hospital with covid-19 in United Kingdom: prospective multicentre observational cohort study. BMJ 2020; 370: m3249.

3. Parri N, Magista AM, Marchetti F, et al. Characteristic of COVID-19 infection in pediatric patients: early findings from two Italian Pediatric Research Networks. European journal of pediatrics 2020; 179(8): 1315–23.

4. Gotzinger F, Santiago-Garcia B, Noguera-Julian A, et al. COVID-19 in children and adolescents in Europe: a multinational, multicentre cohort study. Lancet Child Adolesc Health 2020; 4(9): 653–61.

5. Munro APS, Faust SN. COVID-19 in children: current evidence and key questions. Current opinion in infectious diseases 2020; 33(6): 540–7.

6. Feldstein LR, Rose EB, Randolph AG. Multisystem Inflammatory Syndrome in Children in the United States. Reply. The New England journal of medicine 2020; 383(18): 1794–5.

7. Whittaker E, Bamford A, Kenny J, et al. Clinical Characteristics of 58 Children With a Pediatric Inflammatory Multisystem Syndrome Temporally Associated With SARS-CoV-2. JAMA : the journal of the American Medical Association 2020; 324(3): 259–69.

8. Theoretical Biology Institute for Integrative Biology ETH Zurich. Covid Dashboard: SARS-CoV-2 Variants of Concern in Switzerland (B.1.1.7 Variant - International Comparison). https://cevo-public.github.io/Quantification-of-the-spread-of-a-SARS-CoV-2-variant/ (accessed 15/08/2021).

9. cov-lineages.org. Lineage B.1.1.7. https://cov-lineages.org/lineage.html?lineage=B.1.1.7 (accessed 23/08/2021).

10. Supasa P, Zhou D, Dejnirattisai W, et al. Reduced neutralization of SARS-CoV-2 B.1.1.7 variant by convalescent and vaccine sera. Cell 2021; 184(8): 2201–11 e7.

11. Davies NG, Abbott S, Barnard RC, et al. Estimated transmissibility and impact of SARS-CoV-2 lineage B.1.1.7 in England. Science 2021; 372(6538).

12. Challen R, Brooks-Pollock E, Read JM, Dyson L, Tsaneva-Atanasova K, Danon L. Risk of mortality in patients infected with SARS-CoV-2 variant of concern 202012/1: matched cohort study. BMJ 2021; 372: n579.

13. Reuters. UK coronavirus variant may be more able to infect children: scientists. 21/12/2020. https://www.reuters.com/article/us-health-coronavirus-variant-children-idUKKBN28V2F5 (accessed 15/08/2021).

14. ISARIC 4C (Coronavirus Clinical Characterisation Consortium). Site set-up. https://isaric4c.net/protocols/ (accessed 15/8/2021).

15. Docherty AB, Harrison EM, Green CA, et al. Features of 20 133 UK patients in hospital with covid-19 using the ISARIC WHO Clinical Characterisation Protocol: prospective observational cohort study. BMJ 2020; 369: m1985.

16. National Institute for Health and Clinical Excellence. COVID-19 rapid guideline: children and young people who are immunocompromised NICE guideline [NG174]. Updated 14/08/2020. https://www.nice.org.uk/guidance/ng174?fromsource=mas (accessed 15/08/2021).

17. Recovery Collaborative Group, Horby P, Lim WS, et al. Dexamethasone in Hospitalized Patients with Covid-19. The New England journal of medicine 2021; 384(8): 693–704.

18. Nyberg T, Twohig KA, Harris RJ, et al. Risk of hospital admission for patients with SARS-CoV-2 variant B.1.1.7: cohort analysis. BMJ 2021; 373: n1412.

19. Frampton D, Rampling T, Cross A, et al. Genomic characteristics and clinical effect of the emergent SARS-CoV-2 B.1.1.7 lineage in London, UK: a whole-genome sequencing and hospital-based cohort study. The Lancet infectious diseases 2021.

20. New and Emerging Respiratory Virus Threats Advisory Group (NERVTAG). NERVTAG: Update note on B.1.1.7 severity, 11 February 2021. https://www.gov.uk/government/publications/nervtag-update-note-on-b117-severity-11-february-2021 (accessed 15/08/2021).

21. Brookman S, Cook J, Zucherman M, Broughton S, Harman K, Gupta A. Effect of the new SARS-CoV-2 variant B.1.1.7 on children and young people. Lancet Child Adolesc Health 2021; 5(4): e9–e10.

22. Harwood R, Yan H, Da Camara NT, et al. Which children and young people are at higher risk of severe disease and death after SARS-CoV-2 infection: a systematic review and individual patient meta-analysis. medRxiv 2021: 2021.06.30.21259763.

23. Ward JL, Harwood R, Smith C, et al. Risk factors for intensive care admission and death amongst children and young people admitted to hospital with COVID-19 and PIMS-TS in England during the first pandemic year. medRxiv 2021: 2021.07.01.21259785.

24. Kushner LE, Schroeder AR, Kim J, Mathew R. “For COVID” or “With COVID”: Classification of SARS-CoV-2 Hospitalizations in Children. Hosp Pediatr 2021.

25. Webb NE, Osburn TS. Characteristics of Hospitalized Children Positive for SARS-CoV-2: Experience of a Large Center. Hosp Pediatr 2021; 11(8): e133–e41.

26. Public Health England. COVID-19: infection prevention and control (IPC) Guidance on infection prevention and control for COVID-19 sustained community transmission is occurring across the UK. Updated 10/08/2021. https://www.gov.uk/government/publications/wuhan-novel-coronavirus-infection-prevention-and-control (accessed 23/08/2021).

## References

1. Health Improvement Scotland. Paediatric Early Warning Score (PEWS) Charts. https://ihub.scot/improvement-programmes/scottish-patient-safety-programme-spsp/spsp-programmes-of-work/maternity-and-children-quality-improvement-collaborative-mcqic/paediatric-care/pews/ (accessed 25/08/2021).

3. Public Health England. COVID-19 vaccine surveillance report: Week 20. May 2021. https://www.gov.uk/government/publications/covid-19-vaccine-surveillance-report (accessed 25/08/2021).

4. NHS England. COVID-19 Hospital Activity. https://www.england.nhs.uk/statistics/statistical-work-areas/covid-19-hospital-activity/ (accessed 25/08/2021).

